# Systematic single-variant and gene-based association testing of thousands of phenotypes in 426,370 UK Biobank exomes

**DOI:** 10.1101/2021.06.19.21259117

**Authors:** Konrad J. Karczewski, Matthew Solomonson, Katherine R. Chao, Julia K. Goodrich, Grace Tiao, Wenhan Lu, Bridget M. Riley-Gillis, Ellen A. Tsai, Hye In Kim, Xiuwen Zheng, Fedik Rahimov, Sahar Esmaeeli, A. Jason Grundstad, Mark Reppell, Jeff Waring, Howard Jacob, David Sexton, Paola G. Bronson, Xing Chen, Xinli Hu, Jacqueline I. Goldstein, Daniel King, Christopher Vittal, Timothy Poterba, Duncan S. Palmer, Claire Churchhouse, Daniel P. Howrigan, Wei Zhou, Nicholas A. Watts, Kevin Nguyen, Huy Nguyen, Cara Mason, Christopher Farnham, Charlotte Tolonen, Laura D. Gauthier, Namrata Gupta, Daniel G. MacArthur, Heidi L. Rehm, Cotton Seed, Anthony A. Philippakis, Mark J. Daly, J. Wade Davis, Heiko Runz, Melissa R. Miller, Benjamin M. Neale

## Abstract

Genome-wide association studies have successfully discovered thousands of common variants associated with human diseases and traits, but the landscape of rare variation in human disease has not been explored at scale. Exome sequencing studies of population biobanks provide an opportunity to systematically evaluate the impact of rare coding variation across a wide range of phenotypes to discover genes and allelic series relevant to human health and disease. Here, we present results from systematic association analyses of 4,529 phenotypes using single-variant and gene tests of 426,370 individuals in the UK Biobank with exome sequence data. We find that the discovery of genetic associations is tightly linked to frequency as well as correlated with metrics of deleteriousness and natural selection. We highlight biological findings elucidated by these data and release the dataset as a public resource alongside the Genebass browser for rapidly exploring rare variant association results.

## Introduction

Coding variation has been the most readily interpretable class of genomic variation since the development of the gene model and mapping of the human genome. As such, it has facilitated the mapping and interpretation of variants with immediate clinical importance such as the American College of Medical Genetics actionable variant list (Kalia et al., 2017). More recently, exome sequencing has yielded the discovery of specific causal variants for hundreds of rare diseases, particularly dominant acting *de novo* variants for severe diseases (Bamshad et al., 2019).

As the sample sizes of exome sequencing datasets continue to grow, so do the opportunities to identify associations between rare variants and phenotypes (both complex traits and diseases). In complex diseases, identifying causal genetic factors for a given disease can provide direct insight into the potential for therapeutic avenues. For instance, gain-of-function variants in *PCSK9* have been demonstrated to increase LDL levels and thus risk for cardiovascular disease (Abifadel et al., 2003). Accordingly, loss-of-function (LoF) variants are protective for cardiovascular disease (Cohen et al., 2006), and less than 15 years after the discovery of this effect, therapeutic approaches to inhibit PCSK9 have been brought to market (Sabatine et al., 2017).

Deeply phenotyped biobanks present a unique opportunity to simultaneously analyze multiple diseases and traits within a single cohort, enabling the discovery of new disease genes with therapeutic potential at a large scale, such as the identification of rare variants in *ANGPTL7* that protect against glaucoma (Tanigawa et al., 2020). The UK Biobank is a collection of approximately 500,000 participants with standardized, detailed phenotypic data (Bycroft et al., 2018) on which GWAS have been run extensively. The UKB Exome Sequencing Consortium, a partnership between the UKB and 8 biopharma companies, generated exome sequences for this cohort (Szustakowski et al., 2021), and recent studies have used the exome sequence data to explore various aspects of rare variant associations, including novel biological signals for type 2 diabetes (Deaton et al., 2021), cardiometabolic traits (Jurgens et al., 2022), as well as cross-phenotype analyses that identify new hits for a variety of traits (Backman et al., 2021; Sun et al., 2022; Wang et al., 2021). Here, we describe results from a systematic, large-scale rare variant association analysis of 4,529 phenotypes, release these full sets of summary statistics in a results browser, and explore the role of natural selection and allele frequency on rare variant associations.

### Generating high-quality exome data for rare variant associations

We built an end-to-end pipeline for read mapping, processing, joint variant calling, quality control (QC), and mixed model association analysis, and applied this pipeline to 454,697 individuals with exome sequence data from the UK Biobank. The read mapping and processing pipeline adopted the GATK Best Practices pipeline (GRCh38), and the resulting variants (gVCF files) were joint-called using a scalable implementation in Hail (Supplementary Information; Fig. S1) (Hail Team, 2020). We processed a set of 4,529 phenotypes including 1,233 quantitative traits as well as 3,296 binary traits with at least 200 cases, which included 725 disease endpoints based on ICD-10 codes (Fig. S2).

After performing QC in a similar but augmented (e.g. array concordance; see Supplementary Information) manner as for the Genome Aggregation Database (gnomAD) (Karczewski et al., 2020), we generated a high-quality dataset of 450,953 individuals (Figs. S3 to S5; table S1) including related individuals. This included 426,370 individuals of European ancestry in which we find 23,880,790 high-quality variants (Fig. S6). For each of 19,407 protein-coding genes, we considered up to four functional annotation categories: predicted LoF (pLoF), missense (including low-confidence pLoF variants and in-frame indels), synonymous, and the combination pLoF or missense group, resulting in 8,074,878 variants and 75,767 groups for association testing (i.e., one group per gene and functional annotation category).

### Creating a high-quality set of rare variant associations

We performed group tests using the mixed model framework SAIGE-GENE (Zhou et al., 2020), which includes single-variant tests and gene-based burden (mean), SKAT (variance), and SKAT-O (hybrid variance/mean) tests (Fig. S7). In total, we performed up to 8,074,878 single-variant tests and 75,767 group tests for each of 4,529 phenotypes (Fig. 1). Additionally, we generated 314 heritable random phenotypes to test the asymptotic properties of the mixed-model association testing framework (Figs. S8 to S9), and to determine empirical p-value thresholds for Type I error control. Based on this analysis, for each phenotype, in addition to QC criteria defined below, we consider genome-wide p-value thresholds of 2.5 × 10^−7^ for SKAT-O tests, 6.7 × 10^−7^ for burden tests, and 8 × 10^−9^ for single-variant tests (see Supplementary Information; Fig. S10), corresponding to approximately 0.05 expected false positives per phenotype.

**Figure 1.**
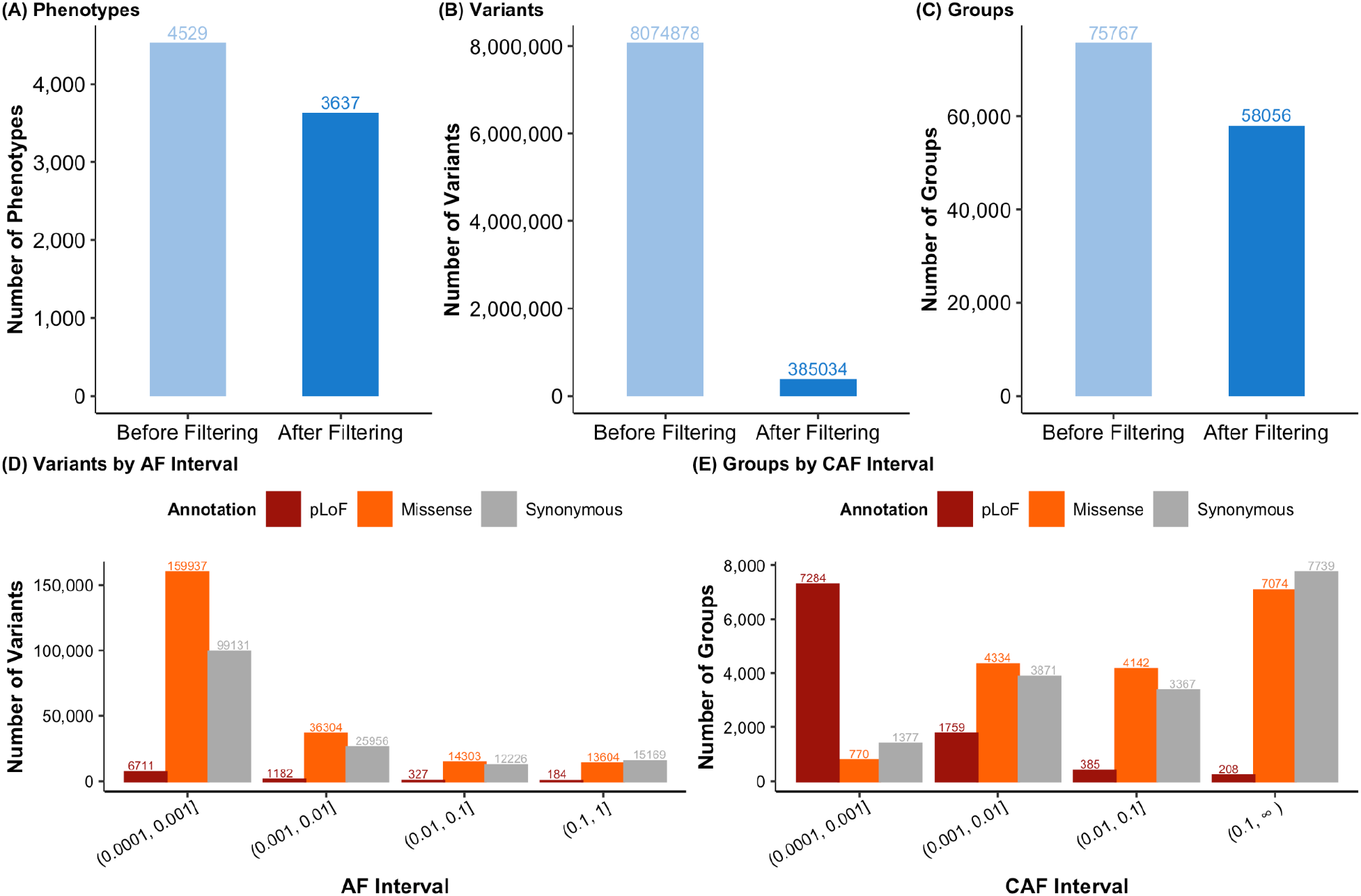
Quality control (QC) of rare variant association tests. The number of phenotypes (**A**), variants (**B**), and groups (i.e., gene-annotation pairs; **C**) before and after QC. After QC, the number of variants (**D**) and genes (**E**) are broken down by annotation and frequency bin (alternate allele frequency [AF] for variants, cumulative allele frequency [CAF] for genes).

We performed extensive QC on these summary statistics (Fig. 1; Table S2; Supplementary Information), including a minimum of two variants per group test, a minimum coverage of 20X, a minimum expected allele count (frequency × n_cases) of 50 for the summary statistics, respectively, as well as genomic control (lambda GC) for each phenotype and each gene (Figs. S11 to S15). Further, we pruned to a set of 3,637 high-quality independent phenotypes encompassing 677 continuous traits and 2,960 binary traits, including 690 ICD codes (Figs. 1A, S16; Table S2). We confirmed the robustness of our results by comparing them to a previous large-scale study of height (Tables S3 to S5, Fig. S17) and red blood cell phenotypes (Table S6), for which our analysis replicates the majority of associations with consistent direction of effect (Hu et al., 2021; Marouli et al., 2017).

We filtered to 385,034 variants, including 8,404 pLoF variants, 224,148 missense variants, and 152,482 synonymous variants with at least one phenotype having expected allele count (cohort frequency × n_cases) over 50 (Fig. 1B). For group tests, we filter to a high-quality set of 58,056 gene tests with at least 20X coverage (Fig. S13) and at least one phenotype with expected allele count ≥ 50 for pLoF (N=9,636 genes), missense (N=16,320), synonymous (N=16,354), and pLoF or missense (N=15,746 genes) (Fig. 1C).

Using these criteria, we identified a total of 71,648 and 6,991 associations meeting our p-value threshold with a mean of 19.7 and 1.9 associations per phenotype, for single-variant tests and group tests, respectively (disease results shown in Fig. 2A-B). Comparing the group test results to single-variant association test results, we find that single-variant tests identify more significant associations than group tests, as these are largely from common variants that are excluded from the group tests. However, we also find 2,237 associations (on average 0.62 per phenotype) from group tests where no single-variant association reached our p-value threshold for any single variant in the corresponding gene (Fig. 2C). Further, most associations arise from missense and synonymous variants, as expected from their greater numbers in the exome, particularly from single-variant associations. However, pLoF variants exhibit relatively more associations in group tests, which is consistent with these variants being individually rare, but directionally consistent, resulting in increased power in a group test (Fig. 2D). In combined tests of pLoF and missense variants, we find an additional 267 associations among burden tests (245 for SKAT-O) that are significant for the combined test but not missense or pLoF tests alone.

**Figure 2.**
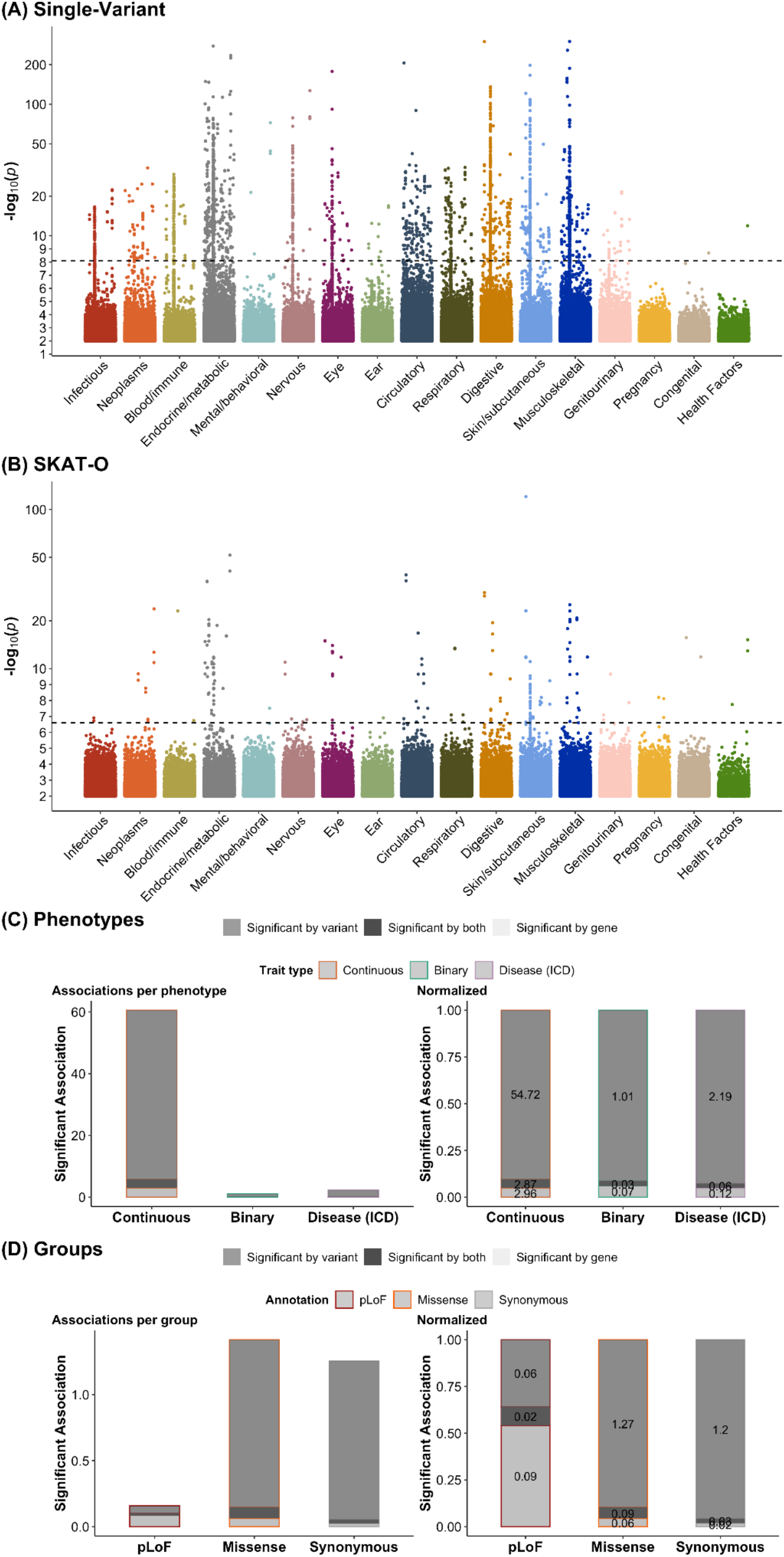
Rare variant association testing is enhanced by group tests. (**A-B)**, For each ICD chapter, we show a Manhattan plot, depicting the distribution of p-values for all single-variant (**A**) and SKAT-O gene-based (**B**) associations, where for each variant/gene, the minimum p-value across phenotypes within each category is shown. (**C-D**): The number of gene-level associations per phenotype is shown as a barplot, broken down by trait type (left) and normalized within each trait type (right), separated by phenotype category (**C**) or functional annotation (**D**). The single-variant tests are grouped into genes where at least one associated variant is necessary to be “Significant by variant” which is shown alongside group tests (“Significant by gene”) as well as genes where an association is found both for group and single-variant tests.

### Displaying rare variant associations

The utility of human genetic variation datasets are substantially enhanced when made accessible in the form of online portals that enable non-technical domain experts to quickly browse, interpret, and export results for downstream follow-up (Karczewski et al., 2017). We extended our gnomAD browser toolkit to create the genebass (gene-biobank association summary statistics) browser (https://genebass.org), a new, highly interactive tool for exploring large numbers of gene-based PheWAS analysis results. This resource provides users with direct access to all 4,529 phenotypes, serving up 993,280,477 gene-level association statistics (across 19,407 genes, 4 annotation sets, and 3 burden tests) and 28,158,190,538 single variant association statistics across 8,074,878 exome variants. For completeness, the released dataset includes all association statistics, including pre-QC data, but we provide functionality to filter to only the highest quality data presented herein. Our web application features a novel layout and navigational scheme for rapidly browsing phenome-wide associations by integrating results across genes and variants. Customizable controls, plots, and tables enable flexible filtering and visualization of phenotypes, genes, and variants of interest; results can be exported for downstream analyses; and variant associations across traits can be compared to inform pathways associated with complex traits and develop therapeutic hypotheses (see Supplementary Information).

### Frequency and selection affect the landscape of rare variant associations

The relationship between natural selection, allele frequency, effect size, and power for discovery is a major complexity in the analysis and interpretation of association statistics, particularly from rare variants. The power to detect association is proportional to the variance explained of a biallelic variant (Sham et al., 2000). Specifically, for a continuous trait the variance explained of a biallelic variant that is purely additive is 2pqa^2^ where p is the allele frequency, q = 1-p and a is the allelic effect of the variant. Thus, for a fixed effect size, a more common variant will capture more variance and by extension show stronger association.

However, the process of negative selection will tend to decrease the frequency of functional damaging variants, suggesting that variants with large effect sizes are more likely to be rare. Indeed, partitioned heritability analyses for common variants support the presence of these countervailing forces, as comparatively lower frequency variants have larger absolute effect sizes but this growth in effect size is slower than the loss in variance explained from their lower frequency (Gazal et al., 2017). In evaluating the landscape of rare variant association, we observe a similar pattern with increasing proportion of variants associated with at least one phenotype as frequency increases (Fig. 3A). However, within each frequency category, we observe the effect of functional annotation, a known correlate for deleteriousness, on the association statistics.

**Figure 3.**
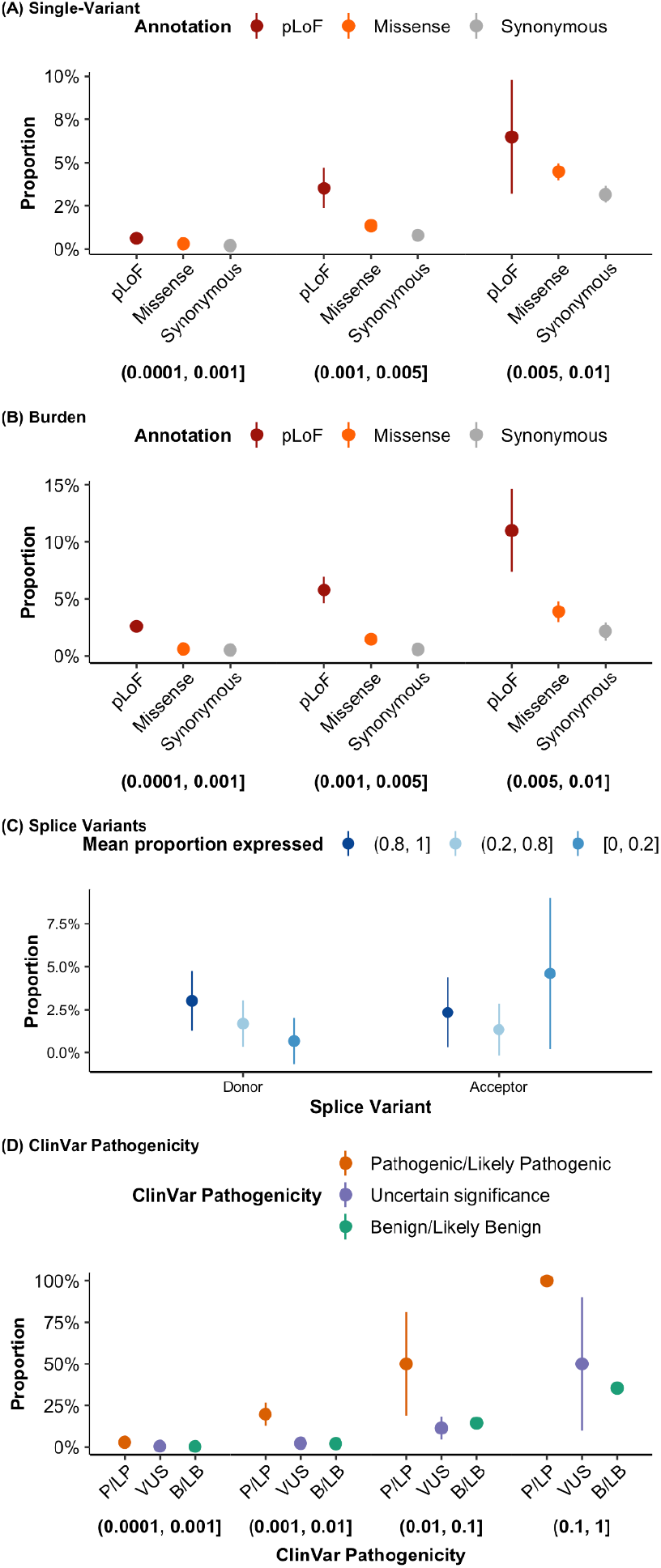
The influence of variant allele frequency and functional annotation in exome association testing. The proportion of single variants (**A**) and genes (**B**) with at least one significant hit is shown broken down by allele frequency category (**A**) or cumulative allele frequency category (**B**), each shown below the plot, broken down by functional annotation. This metric is also plotted by the proportion expressed across transcripts for splice variants (**C**), and ClinVar pathogenicity status (**D**).

Comparing the number of associations by variant annotation in each allele frequency category, we find that pLoF variants have a larger number of associations than missense variants, followed by synonymous variants for single-variant tests (Fig. 3A) as well as group tests (Fig. 3B). For common variants (>1%), we observe further increases in associations due to power, but with attenuated associations for pLoF variants, likely due to an increased rate of artifacts at common pLoF variants (MacArthur and Tyler-Smith, 2010) (Fig. S18). Within missense variants, variant deleteriousness as predicted by PolyPhen2 (Adzhubei et al., 2010) is correlated with the number of associations meeting our p-value threshold (Fig. S18). For splice donor variants, we find a correlation between the proportion expressed across transcripts (pext) (Cummings et al., 2020) and the number of associations (Fig. 3C). Additionally, the pathogenicity level of ClinVar variants is correlated with phenotypic association (Fig. 3D).

### Gene function influences association statistics

We examined the phenotypic impact of gene categories previously known to have functional relevance and/or a role in disease. In particular, we find that 470 genes previously implicated in developmental delay (Kaplanis et al., 2020) are more likely to be associated with a phenotype in the UK Biobank (Fisher’s exact p = 3.6 × 10^−4^, OR = 3.50; Fig. 4). Further, we observe a correlation between selection against pLoFs in a gene and the phenotypic impact of pLoFs in that gene: specifically, constrained genes (i.e., those in the highest decile of LoF observed/expected upper bound fraction [LOEUF], a metric of LoF intolerance) are more likely to be associated with a phenotype (9.14%) than a frequency-matched set of genes in the genome (2.12%; Fisher’s exact p = 6.1 × 10^−14^, OR = 4.65; Fig. 4). Similarly, genes with known autosomal dominant and autosomal recessive diseases, as well as genes with previously established hits in the GWAS catalog and FDA approved drug targets, show an increased phenotypic impact of pLoFs and missense variants.

**Figure 4.**
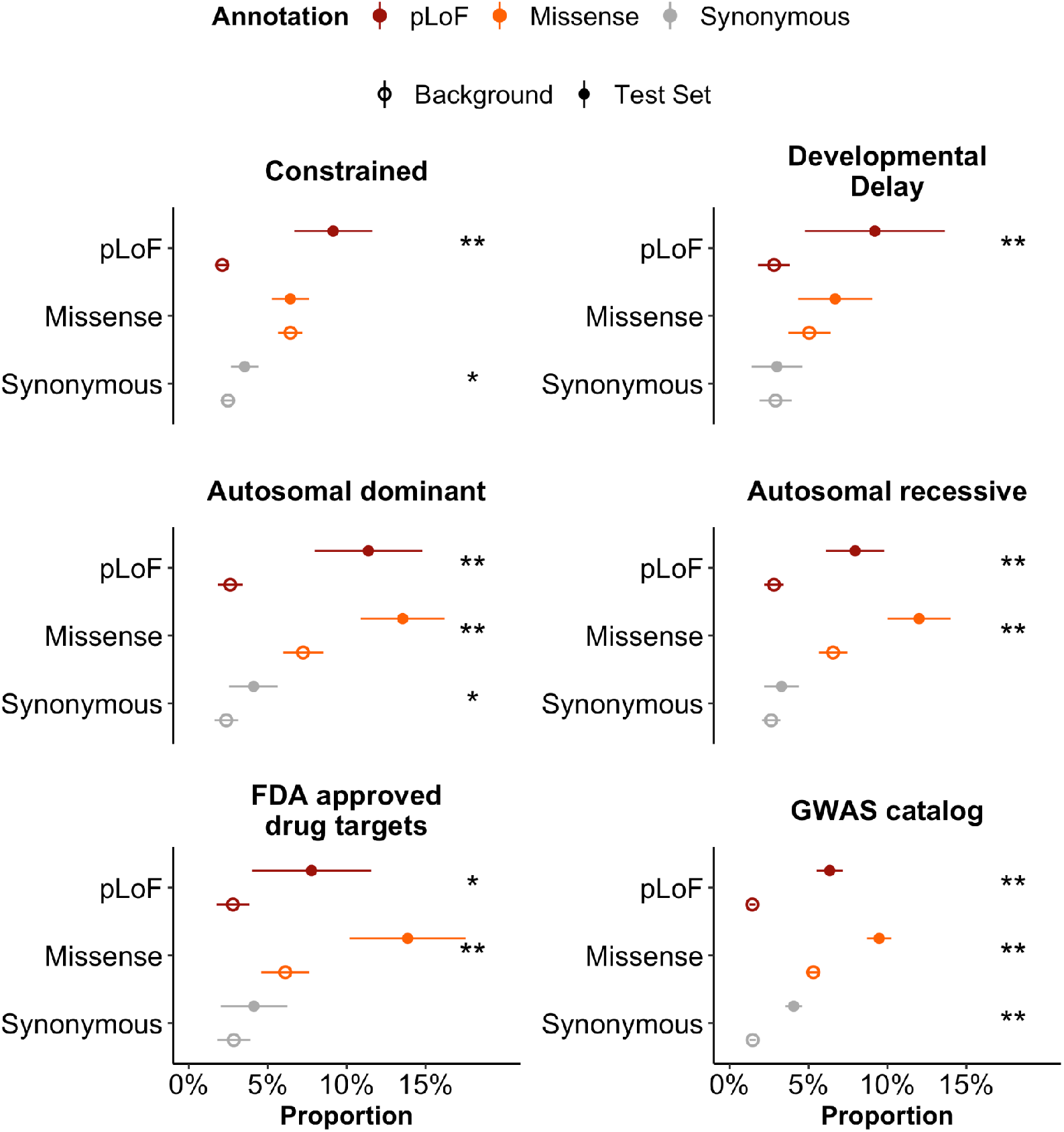
The effect of gene function on the landscape of rare variant associations. The proportion of gene-annotation pairs with at least one association (SKAT-O p < 2.5 × 10^−7^) is shown for a number of gene categories, each compared to a background set of genes matched on cumulative allele frequency. Error bars represent 95% confidence intervals. Asterisks denote a significant difference between the background set and test set (* and ** indicate p < 0.05 and p < 0.001, respectively).

### Biological insights from rare variant association results

The biological information encapsulated in this dataset is extremely high-dimensional, and we release the full dataset of results for the benefit of the community. Here, we highlight a set of known and putative associations as examples of the power of this dataset. First, we recapitulate many known associations from previous studies, including associations between *PCSK9* and LDL cholesterol (pLoF burden p = 3.5 × 10^−132^), *COL1A1* and bone density (pLoF burden p = 2 × 10^−9^) (Mann et al., 2001), *KLF1* and several red blood cell traits (pLoF burden < 2 × 10^−12^) (Perkins et al., 2016), and *LRP5* (Wnt coreceptor) and bone density and osteoporosis phenotypes (pLoF burden < 5 × 10^−7^) (Baron and Rawadi, 2007).

We highlight novel biological signals identified in the exome dataset, enabled by the Genebass browser. In particular, we find an association between predicted loss-of-function of *SCRIB* and white matter integrity of tapetum (Fig. 5). Notably, this association is not significant at any single pLoF variant, but when aggregated into a SKAT-O or burden group test, the overall ablation of the transcript is associated at a p-value of 6 × 10^−15^ (Fig. 5A). This provides additional context to a signal observed in a recent GWAS of white matter integrity (Zhao et al., 2020) averaged across regions of the brain, as well as in the body of corpus callosum (Fig. 5B). To our knowledge, this gene has not been associated in previous genome-wide association studies, although it is a constrained gene (pLI = 0.93) that shows evidence for neural tube defects in mice (Murdoch et al., 2003) with ultra-rare occurrences in humans (Lei et al., 2013).

**Figure 5.**
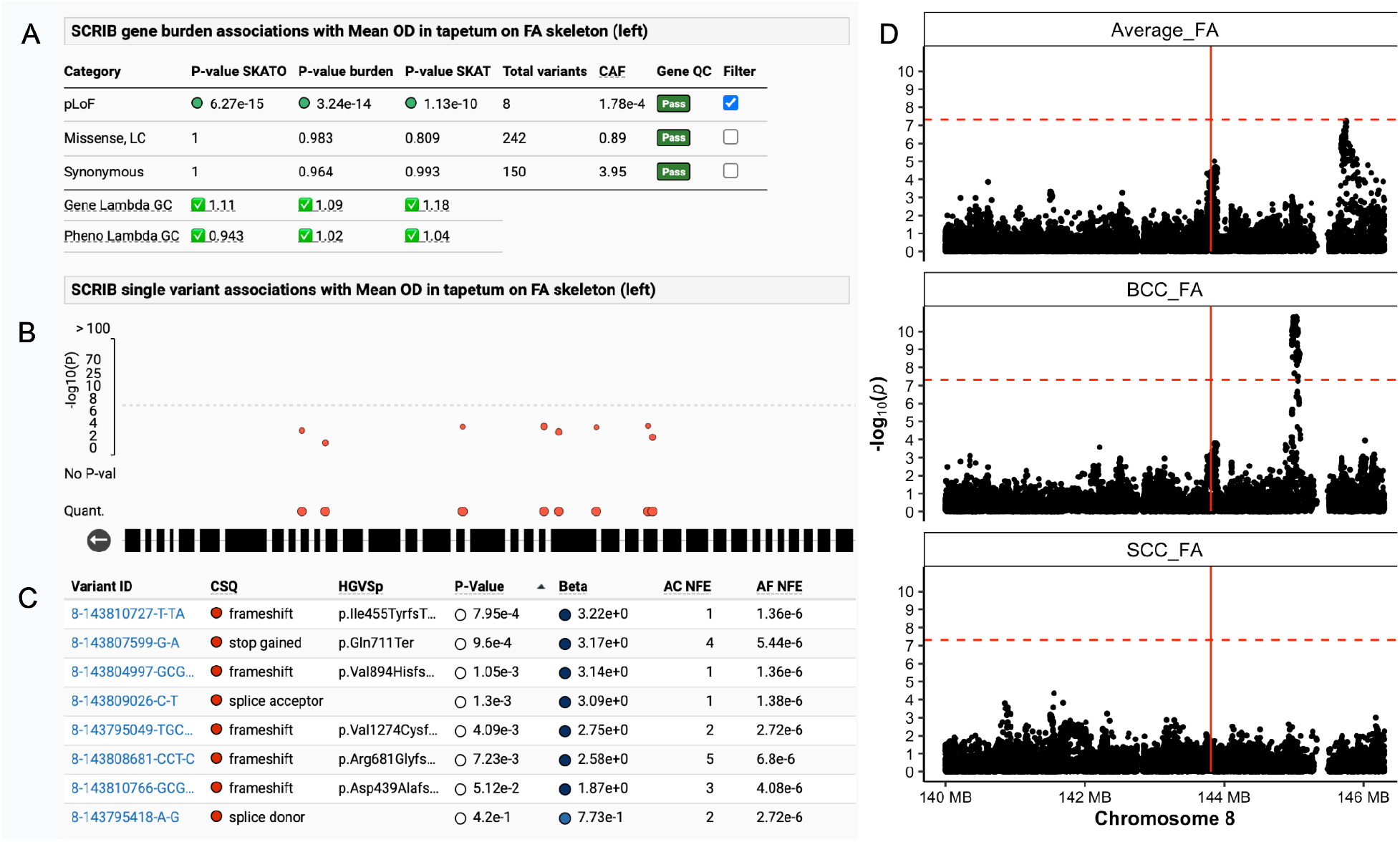
Refined association between *SCRIB* and white matter integrity of tapetum. The Genebass browser provides views of the full dataset, including all quality control metrics and association statistics. (**A**): The summary of association information between pLoF variants in *SCRIB* with mean OD (orientation dispersion index) in tapetum on FA (fractional anisotropy) skeleton (from dMRI data). (**B**): A rare variant Manhattan plot of 8 rare pLoF variants is shown. (**C**): Details for the component variants are shown in a table, including their functional consequence (CSQ), a detailed protein-coding annotation (HGVSp), the association p-value and beta, as well as frequency information (AC: allele count, Hom: number of homozygotes, AN: allele number, AF: allele frequency). Each component pLoF variant in scrib has a positive beta value and in aggregate, these variants show an association at p = 6 × 10^−15^ (**A**). (**D**): A Manhattan plot of a previous GWAS (Zhao et al., 2020) of FA averaged across brain regions (top), body of corpus callosum (middle), and splenium of corpus callosum (bottom). Horizontal dashed line indicates a GWAS genome-wide significance threshold (5 × 10^−8^), and vertical line indicates the location of *SCRIB*.

## Discussion

We have generated rare variant association analysis summary statistics for 4,529 phenotypes and made these data available to the public, via bulk data downloads as well as a public-facing browser (https://genebass.org). We explore aspects of this resource relating natural selection, allele frequency, and genetic discovery, and we highlight a novel association between *SCRIB* and a brain imaging trait. Future work will be needed to fully assess the contribution of rare variants to the heritability of common diseases, as well as the extent and role of pleiotropy among rare variants.

### Limitations of the study

There are a number of limitations to our analysis. Although we performed extensive QC to improve the reliability of these results, we urge caution in interpreting association results, particularly for the rarest binary traits (prevalence < 10^−4^) and ultra-rare variants (frequency < 10^−4^) as the asymptotic properties of the association tests may not be met. For the rarest outcomes, increasing the number of cases is essential to properly evaluate the impact of rare coding variation across genes. Alternatively, other statistical methods such as Firth regression may be better suited to such traits. For pLoF variants, the median cumulative allele frequency across genes is approximately 1.5 × 10^−4^, suggesting that group tests at current sample sizes are only powered to detect individual gene effects for quantitative traits that capture at least 0.02% of variance, as well as diseases and traits that have a high prevalence (well above 10%; Fig. S10). These considerations are underscored by the apparently poor asymptotic properties of the mixed-model tests for rarer binary traits, as the lambda GC for these tests decreases precipitously (Fig. S9). Nonetheless, global biological trends are apparent, such as the relative ordering of functional impact (pLoF > missense > synonymous; Fig. 3), highlighting that the ability to accurately annotate variants with the functional consequences on a gene is critical to powering discovery in rare variant analysis. Further, measures of natural selection at the gene level continue to highlight that certain classes of genes, such as LoF-intolerant genes, are clearly enriched for phenotypic associations.

Finally, these association analyses were only performed for individuals of European ancestry, the largest group in the dataset. Notably, these analyses only interrogate a slice of human genetic diversity, and expanding to additional ancestries has been shown to increase power and resolution for genetic discovery (Majara et al., 2021; Morales et al., 2018; Sakaue et al., 2020); however, as the sample sizes of non-European individuals in the UK Biobank are very limited, these analyses would be underpowered for most binary traits including many disease outcomes. Concentrated efforts in building large biobanks with diverse participants will be required to overcome these limitations and provide more insight into the contribution of rare variants to common disease etiology.

### Biobank teams

#### Abbvie

- <u>Steering team:</u> Jeff Waring, Howard Jacob, J. Wade Davis
- <u>Data management team</u>: A. Jason Grundstad, Silvia Orozco
- <u>Extended Scientific team</u>: Bridget Riley-Gillis, Sahar Esmaeeli, Fedik Rahimov, Ali Abbasi, Nizar Smaoui, Xiuwen Zheng, Emily King, John Lee, Reza Hammond, Mark Reppell, Hyun Ji Noh

#### Biogen

- <u>Steering team:</u> Ellen Tsai, Christopher D. Whelan, Paola G. Bronson, David Sexton, Sally John, Heiko Runz
- <u>Data management team</u>: Eric Marshall, Mehool Patel, Saranya Duraisamy, Timothy Swan
- <u>Extended Scientific team:</u> Denis Baird, Chia-Yen Chen, Susan Eaton, Jake Gagnon, Feng Gao, Cynthia Gubbels, Yunfeng Huang, Varant Kupelian, Kejie Li, Dawei Liu, Stephanie Loomis, Helen McLaughlin, Adele Mitchell, Nilanjana Sadhu, Benjamin Sun, Ruoyu Tian

#### Pfizer

- <u>Steering team</u>: Hye In Kim, Xinli Hu, Morten Sogaard, A. Katrina Loomis, Eric Fauman, Melissa R. Miller
- <u>Data Management team</u>: Jay Bergeron, Andrew Hill, Juha Sarimaa, Zhan Ye, Xing Chen
- <u>Extended Scientific team</u>: Yi-Pin Lai, Jean-Philippe Fortin, Joanne Berghout, Robert Moccia, Craig L. Hyde

## Data Availability

All data is publicly available at https://genebass.org

## Acknowledgements

We thank Danielle Ciofani and Cathy Marshall for their efforts in launching this project. We thank the participants and leadership of the UK Biobank: this work was done under UK Biobank applications 26041 and 48511.

