## Supplementary Information for "Systematic single-variant and gene-based association testing of thousands of phenotypes in 426,370 UK Biobank exomes"

### UKB Exomes Supplement

**Data processing and joint calling** 4
Data processing 4
Joint calling 4
Representation 5
Fig. S1 | Scalable Variant Call Representation (SVCR) created from two gVCF inputs. 6
Merge algorithm 7
Phenotype data processing 8
Fig. S2 | Phenotype curation pipeline. 9

**Genotype, Sample, and Variant Quality Control** 11
Concordance between arrays and exomes 11
Interval QC 11
Fig. S3 | Histogram showing the percentage of samples meeting 20X mean coverage for each exome capture interval. 12
Sample QC 12
Sex imputation 12
Fig. S4 | Normalized chromosome X ploidy plotted against normalized chromosome Y ploidy and colored by sex karyotype. 13
Hard filters 13
Platform inference 14
Fig. S5 | Platform inference using missingness PCA. 14
Relatedness inference 15
Ancestry imputation 15
Outlier detection 16
Table S1 | Final sample counts passing QC. 16
Variant QC 16
Fig. S6 | Variant QC 19
Genotype QC 19
Annotations 19

**Scaling association testing using Hail Batch** 21
Motivation 21
User interface 22
Fig. S7 | Hail Batch schematic for SAIGE association analysis. 23
Implementation 24

**Association testing and quality control** 26
Association testing framework 26
Random phenotype analysis 27

Fig. S8 | QQ-plots of randomly generated heritable (heritability = 100%) phenotypes for single-variant tests (left) and for group tests (SKAT-O, middle; and burden tests, right). 28
Fig. S9 | Lambda GC by cumulative allele frequency (CAF) by heritability. 29
Fig. S10 | Power for rare variant associations. 30
Expected allele count filters 30
Fig. S11 | Lambda GC for each phenotype vs case count, split by expected AC interval for SKAT-O, SKAT and burden tests. 31
Test calibration filtering 32
Fig. S12 | Lambda GC for each phenotype vs case count for SKAT-O, SKAT and burden tests, before and after filtering out summary statistics with expected AC < 50, number of variants tested < 2, and coverage < 20. 32
Fig. S13 | Coverage vs gene-based lambda GC, for SKAT-O (A) and burden tests (B). 34
Fig. S14 | Lambda GC for each phenotype. 34
Fig. S15 | Lambda GC for each gene. 35
Independent phenotypes 35
Fig. S16 | Independence of phenotypes. 36
Table S2 | QC of summary statistics. 37
Comparison to known hits 38
Table S3 | Comparison to 32 rare (MAF < 1%) variants associated with adult height in GIANT. 38
Table S4 | Comparison to 59 low-frequency (MAF between 1% and 5%) variants associated with adult height in GIANT. 40
Table S5 | Comparison to 10 genes associated with adult height in GIANT. 43
Fig. S17 | Comparison of effect sizes between UK Biobank and GIANT for height. 45
Table S6 | Comparison of 20 associations between missense variants and 7 major red blood cell phenotypes discovered at the genome-wide significant loci of the marginal tests in TOPMed. 45
**Analysis of summary statistics** 47
Fig. S18 | The proportion of genes (A) and variants (B) associated with at least one trait, broken down by functional class. 47
Fig. S19 | The proportion of variants (A) and genes (B) with at least one phenotype reaching our p-value threshold is shown broken down by allele frequency category (A) or cumulative allele frequency category (B) and by functional category. 48
Gene set analyses 49
Developmental delay genes 49
Constrained genes 49
Other gene sets 50
PolyPhen2 predicted variants 50
Fig. S20 | The proportion of variants with at least one association is shown broken down by PolyPhen2 annotation group and allele frequency category. 51
ClinVar variants 51

**Data availability and release** 53
Data availability 53
Code availability 53
Data browser 53
Navigation and workflow 54
Fig. S21 | Overview of the UKBB exome gene browser interface. 55
Exploring associations by gene 55
Exploring associations by phenotype 57
Fig. S22 | Results by phenotype. 57
Comparing single variant associations across phenotypes 58
Fig. S23 | Multi-phenotype plotting. 58
Fig. S24 | Using hover interactions with the multi-phenotype pivot table. 60
Fig. S25 | Viewing case-control counts and allele frequencies for pLoF variants across traits in a gene. 61
Fig. S26 | Color variants by attribute to uncover patterns in A) consequence, B) p-value, C) beta, D) trait, or E) zygosity. 62
Single variant results 62
Fig. S27 | Single variant page. 63
**References** 64

### Data processing and joint calling

Timothy Poterba, Chris Vittal, Duncan Palmer, Cara Mason, Christopher Farnham, Charlotte Tolonen, Laura D. Gauthier, Konrad J. Karczewski, Namrata Gupta, Claire Churchhouse, Daniel G. MacArthur, Anthony Philippakis, Cotton Seed, Melissa R. Miller, Benjamin M. Neale

**Data processing**

We re-processed CRAM files according to the GATK Best Practices, briefly, aligning reads using BWA-MEM 0.7.15.r1140 and processing reads using Picard and GATK. For each sample, variants were called using the Genome Analysis Toolkit (GATK) 4.0.10.1 with local realignment by HaplotypeCaller in gVCF mode, such that every position in the genome is assigned likelihoods for discovered variants or for the reference. We then post-processed gVCF outputs, further compressing "blocks" of homozygous reference calls to seven GQ bins: 0, 10, 20, 30, 40, 50, and 60. All analyses were performed on Google Cloud Platform.

**Joint calling**

Many technologies used to represent, store, and compute on genomic data do not scale efficiently to datasets of hundreds of thousands of samples with whole exome or genome sequence data. The Hail project addresses these scaling limitations by developing new technology to enable representation and analysis of cohorts with hundreds of thousands of samples. The project VCF (pVCF) representation scales poorly, and one key innovation that allowed efficient analysis of such a large callset is the Scalable Variant Call Representation (SVCR), which takes advantage of reference block compression to guarantee strictly linear asymptotic scaling with the number of stored samples. The SVCR is a conceptual representation for the variant-level data on a sequenced cohort. In the Hail library, we have developed an implementation of an efficient hierarchical merge algorithm to create a dataset in Hail conforming to the SVCR representation from single-sample gVCFs, and a rudimentary set of functionality for working with this object as an analysis target.

*Representation*

We will briefly describe the representation of the SVCR and where it differs from a pVCF. First, the SVCR contains a row for each locus defined in any input gVCF. While pVCF only includes a row for each variant site, the SVCR contains rows for monomorphic sites where reference blocks begin. The SVCR contains more loci than an equivalent pVCF because while pVCFs only contain loci with genomic polymorphism, the SVCR contains loci where reference blocks begin but with no variation. This difference in locus cardinality shrinks with increasing sample size when rare variants are discovered at most positions in the genome. The alleles field of the SVCR at a given locus contains the set of all unique alternate alleles observed in any input gVCF for that locus (Fig. S1).

Sample 1 GVCF

**a.**

| CHR | POS | Ref | Alt | Sample1 |
|---|---|---|---|---|
| chr1 | 3330 | C | <*NON_REF*> | GT=0/0, DP=10, GQ=30, END=3330 |
| chr1 | 3331 | T | G, <*NON_REF*> | GT=0/1, DP=10, GQ=99, AD=[6, 4], PL=[100, 0, 105] |
| chr1 | 3332 | C | <*NON_REF*> | GT=0/0, DP=11, GQ=30, END=3349 |
| chr1 | 3350 | A | C, <*NON_REF*> | GT=1/1, DP=15, GQ=55, AD=[0, 15], PL=[1002, 55, 0] |

Sample 2 GVCF

**b.**

| CHR | POS | Ref | Alt | Sample2 |
|---|---|---|---|---|
| chr1 | 3330 | C | <*NON_REF*> | GT=0/0, DP=9, GQ=30, END=3334 |
| chr1 | 3335 | G | C, <*NON_REF*> | GT=1/1, DP=7, GQ=22 AD=[0, 7], PL=[154, 22, 0] |
| chr1 | 3336 | T | <*NON_REF*> | GT=0/0, DP=10, GQ=30, END=3349 |
| chr1 | 3350 | A | T, <*NON_REF*> | GT=0/1, DP=12, GQ=99 AD=[7, 5], PL=[154, 0, 102] |

Merged SVCR

**c.**

| CHR | POS | Ref | Alt | Sample1 | Sample2 |
|---|---|---|---|---|---|
| chr1 | 3330 | C | <*NON_REF*> | LGT=0/0, DP=10, GQ=30, END=3330 | LGT=0/0, DP=9, GQ=30, END=3334 |
| chr1 | 3331 | T | G, <*NON_REF*> | LA=[0, 1], LGT=0/1, DP=10, GQ=99, LAD=[6, 4], LPL=[100, 0, 105] | |
| chr1 | 3332 | C | <*NON_REF*> | LGT=0/0, DP=11, GQ=30, END=3349 | |
| chr1 | 3335 | G | C, <*NON_REF*> | | LA=[0, 1], LGT=1/1, DP=7, GQ=22 LAD=[0, 7], LPL=[154, 22, 0] |
| chr1 | 3336 | T | <*NON_REF*> | | LGT=0/0, DP=10, GQ=30, END=3349 |
| chr1 | 3350 | A | C, T, <*NON_REF*> | LA=[0, 1], LGT=1/1, DP=15, GQ=55, LAD=[0, 15], LPL=[1002, 55, 0] | LA=[0, 2], LGT=0/1, DP=12, GQ=99 LAD=[7, 5], LPL=[154, 0, 102] |

Fig. S1 | Scalable Variant Call Representation (SVCR) created from two gVCF inputs. Panels a and b display information contained in gVCFs for two distinct samples in a small genomic window. Panel c represents the merged SVCR, which contains all loci present in either a or b. There is no entry for Sample2 at chr1:3331 because Sample2's gVCF does not contain the locus chr1:3331. The GT field has been renamed to LGT (local GT), and the LA (local alleles) field has been added to record the original alleles in each gVCF, which is important at chr1:3350, a locus where both input samples have a variant call.

An entry of the SVCR matrix corresponding to locus L and sample S is dense (defined) if the gVCF for sample S has a line with locus L, and is sparse (missing) otherwise. If defined, the entry includes a light transformation of all information from the gVCF for sample S at locus L, with the two exceptions of the locus information (chromosome and position) and the alleles (reference and alternates). The chromosome and position are necessarily the same as those constituting L, and instead of including alleles as strings, the SVCR has an additional field, LA, or “local alleles”, which is a list of integers. This list contains the indices in the list of all discovered alleles at the current locus corresponding to alleles that were discovered in the GVCF for sample S. The final transformation is a rename of GT and all fields that are “R-” or “A-” numbered, such as AD and PL, to LGT, LAD, LPL, etc. This rename reflects the change in meaning -- the alleles referred to by these fields are the ones defined in LA, rather than the full list of discovered alleles in the SVCR row.

It is important to note that the SVCR representation is a lossless transformation of gVCFs -- the original gVCFs can be recreated by reversing the transformations defined above.

*Merge algorithm*

We have proposed the SVCR as an efficient conceptual representation for efficiently storing cohort-level data sequencing data. We now describe a hierarchical merge algorithm that lends itself to scalable construction of a SVCR.

First, each input gVCF is converted to a single-sample SVCR. This conversion involves trivial reorganizations and renames of fields (GT to LGT, as described above). The result is a SVCR with the same number of rows as the gVCF and one column, where every entry is defined.

Second, N SVCR are merged together into a new SVCR. First, an outer join on loci is executed for the N SVCRs, producing some number of intermediate rows with a locus (the join key), the alleles for each input SVCR (which may be missing if a SVCR did not contain that locus), and the entries associated with that locus for each SVCR. The merged alleles are computed by taking the set of all unique alleles from all input SVCR matrices, and sorting according to a deterministic string ordering function. The merged entries are computed by taking the full set of entries across join inputs, and updating one field: the values of each LA (local alleles) field must be recomputed to refer to the new, merged alternate alleles.

The Hail framework features a concrete implementation of this hierarchical merge algorithm, repeating the second step in rounds until a single SVCR remains. In practice, this algorithm defaults to a value of 100 for the branch factor parameter, N, which performs well empirically and allows for the creation of a million-sample dataset in three rounds of merging.

*Dense matrix analysis*

The significant scaling advantages of the SVCR representation are not without cost. The sparsity prevents random access to reference block information at any specific variant, because most of the reference blocks spanning that variant will have been defined at earlier loci. Analyses that require access to reference block metadata (such as GQ or DP) can be implemented following a "densification" pass that carries forward an array of genotype values, one for each sample, corresponding to the most recent reference block entry on the same chromosome. The reference blocks in this array can be used to fill sparse entries for each locus in the SVCR (as long as a candidate reference block spans the locus of interest). The necessary dense analysis target can thus be realized at low cost on the fly during analysis pipelines, then discarded. It is not necessary to save the dense matrix to durable storage, which would incur prohibitive cost.

**Phenotype data processing**

To automate the curation and harmonization of the large collection of variable scalings, encodings, and follow-up responses in a coherent manner, we created a modified version of the PHEnome Scan ANalysis Tool (PHESANT), available at https://github.com/astheeggeggs/PHESANT. Unlike the original implementation (Millard et al., 2018), our version does not perform association analyses, but simply generates a collection of re-coded phenotypes. An outline of the PHESANT pipeline and our chosen filters are displayed in Fig. S2. We manually curated the collection of phenotypes for study, which we filter to as part of the pipeline. Re-codings of variables, and inherent orderings of ordinal categorical variables, are defined in the data-coding file, which is available in our GitHub repository.

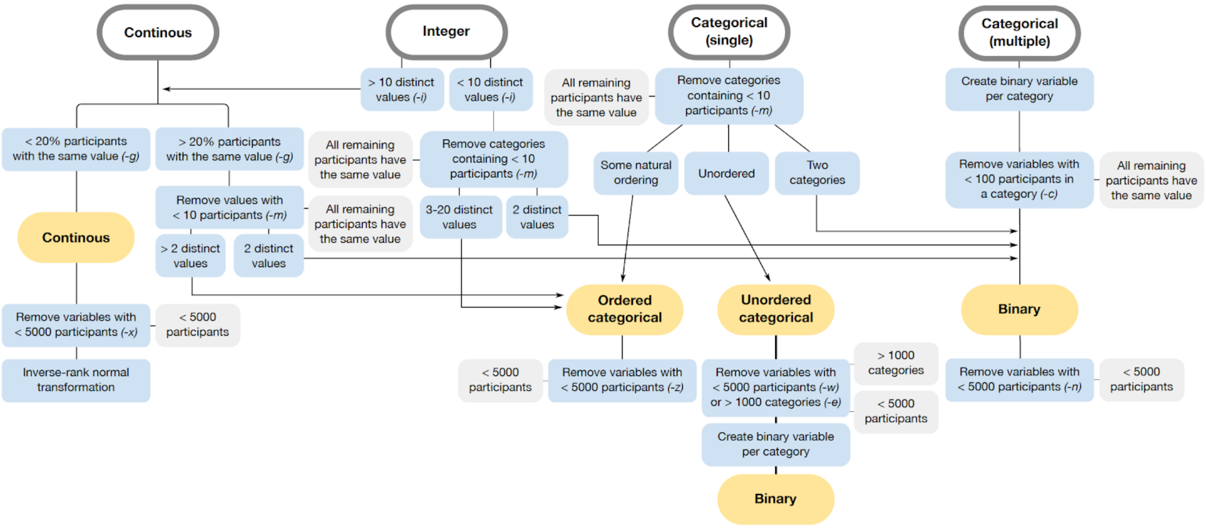


Fig. S2 | Phenotype curation pipeline. Raw phenotype data (gray outlined boxes) are passed to PHESANT, and a collection of filters (blue boxes) are applied. The thresholds shown here are the defaults in our modified version of PHESANT that can be altered in our code as desired using the flags displayed in parentheses. Grey filled boxes display the criteria for removal, and yellow filled boxes show the category of the variable after the rules in the blue boxes have been enforced.

In addition to the inverse-rank normalization applied to the collection of continuous phenotypes, we also retain the raw version of the continuous phenotype, with no transformation applied to the data (though re-codings of the data to guard against spurious results are retained). Following all of these alterations and additions, we run this modified version of PHESANT on the phenotypes in our UK Biobank application using a 200Gb RAM virtual machine on the Google Cloud Platform.

Upon applying the PHESANT pipeline to our selected collection of phenotypes, a small subset of categorical variables remain that should be sex-specific but are not excluded from the both-sexes collection of phenotypes. We manually identified this collection of sex-specific phenotypes and removed them from the both-sex phenotypes before subsequent analysis.

We loaded all phenotype data into a Hail MatrixTable using a custom processing script (https://github.com/Nealelab/ukb_exomes/blob/master/hail/load_phenotype_data.py). Briefly, we parsed the PHESANT output, extracted ICD codes from the "first occurrence" data (which were run as binarized outcomes), parsed some custom phenotypes and covariates, and combined these into a phenotype MatrixTable. The MatrixTable is keyed by trait type (continuous, categorical, and ICD code), phenocode, sex, coding (for categorical traits), and modifier (raw vs inverse-rank normalized for continuous traits). For this supplement, we use the descriptors "categorical" and "binary" interchangeably when describing phenotypes.

### Genotype, Sample, and Variant Quality Control

Julia K. Goodrich, Katherine R. Chao, Laura Gauthier, Konrad J. Karczewski, Benjamin M. Neale, Daniel G. MacArthur, Grace Tiao

We performed quality control (QC) in a similar fashion to the approach used for the Genome Aggregation Database (gnomAD) (Karczewski et al., 2020). Notably, however, we included a number of additional metrics, including concordance between arrays and exomes and interval QC.

**Concordance between arrays and exomes**

We confirmed that all 453,644 samples with available array data had a high proportion of variant concordance between their exome and array data. We filtered the UK Biobank array data to autosomal regions and lifted the data to genome build GRCh38 prior to examining concordance. The minimum proportion concordance of non-reference sample genotypes was 0.96 (mean 0.995).

**Interval QC**

We performed quality controls on intervals targeted by the exome capture before applying sample hard filters. In this interval QC, we investigated how much padding to add around the UK Biobank capture intervals and whether to check coverage across standard Broad exome calling intervals (available on Google Cloud Platform at gs://gcp-public-data--broad-references/hg38/v0/exome_calling_regions.v1.interval_list) as well. We determined that padding more than 50 base pairs into the intron added too much noise and reduced sample call rate and coverage significantly. We also discovered that adding calling intervals unique to the Broad's set of exome targets also reduced call rate and coverage. Therefore, we decided to keep variants only within the 50 base pair padded UK Biobank intervals. For sample QC, we also decided to filter to intervals where 85% of samples had a mean coverage of at least 20X (Fig. S3).

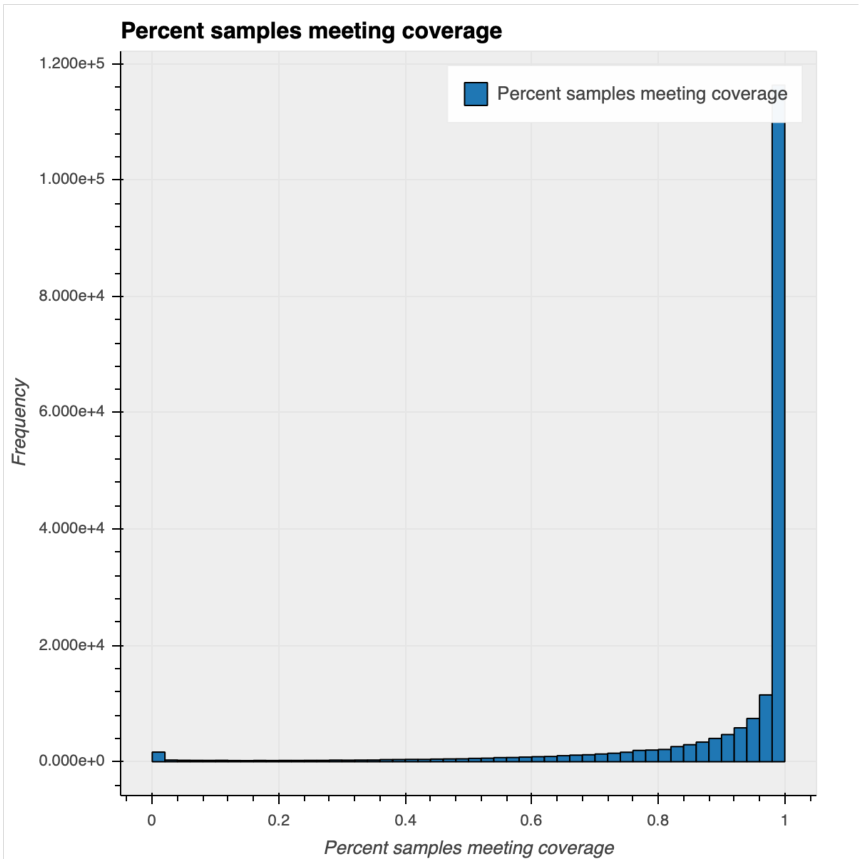


Fig. S3 | Histogram showing the percentage of samples meeting 20X mean coverage for each exome capture interval.

#### Sample QC

##### *Sex imputation*

We used Hail's *impute_sex* method to infer sex using common (allele frequency > 0.1%), non-pseudoautosomal, bi-allelic single nucleotide variants (SNVs) on chromosome X. We then computed non-pseudoautosomal chromosome X and chromosome Y coverage for each sample and normalized these values using sample-specific coverage across chromosome 20. We then checked the distribution of chromosome X and Y ploidies for XX and XY karyotypes to determine each karyotype's ploidy cutoffs (upper cutoff for single X: 1.55, lower cutoff for double X: 1.6, upper cutoff for double X: 2.15, lower cutoff for triple X: 2.2, lower cutoff for single Y: 0.06, upper cutoff for single Y: 1.3, lower cutoff for double Y: 1.4). The adjusted ploidy cutoffs helped us add additional granularity to the inferred sex, differentiating between X0, XX, XXX, XY, XXY, XYY, and XXYY karyotypes (Fig. S4).

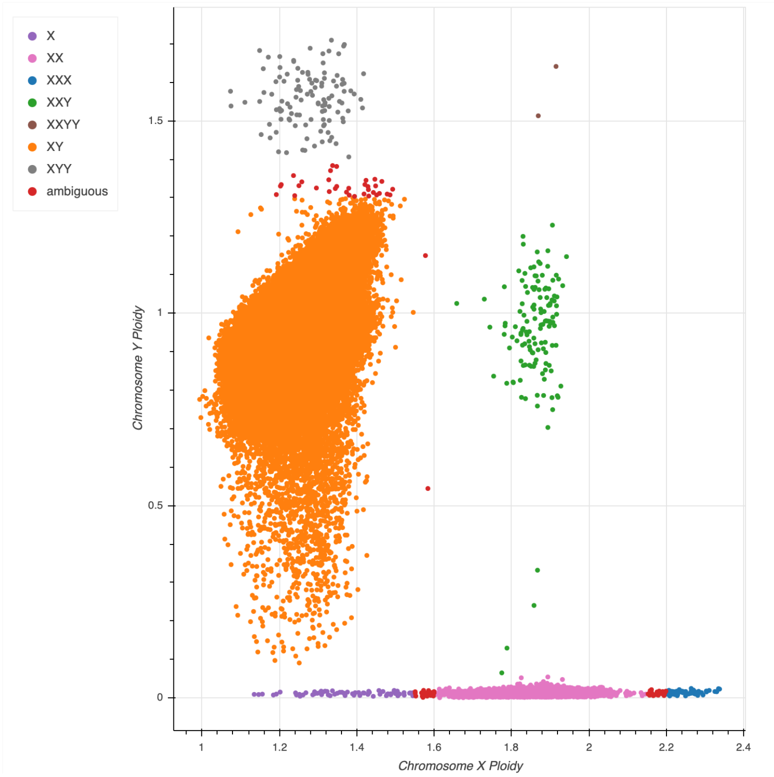


Fig. S4 | Normalized chromosome X ploidy plotted against normalized chromosome Y ploidy and colored by sex karyotype. XY samples are spread out in terms of their normalized chromosome Y coverage. This long tail of samples is likely due to mosaic loss of chromosome Y.

*Hard filters*

We applied three hard filters to the samples: sex imputation filters (removing any samples that were not inferred as XX or XY), a call rate filter (cutoff: 0.99), and a coverage filter (mean coverage cutoff 20X). We excluded hard-filtered samples from platform PCA, relatedness inference, ancestry imputation, and outlier detection so that these low quality samples would not influence our downstream results.

*Platform inference*

Although we do not expect there to be noticeable technical artifacts given that the samples were run on the same platform, we ran a principal component analysis (PCA) on the per-individual per-interval call rate matrix, as previously described (Karczewski et al., 2020), to make sure there were no significant clusters. Our platform PCA picked up a few clusters and showed some differences between the different sample batches on PCs 3 and 4 (Fig. S5). After some investigation, we discovered that the PC4 separated samples due to a common CNV (chr12:9531541-9531809). As PC3 showed some separation by batch, we decided to use batch status as a proxy for platform in further sample QC and a covariate for association analysis.

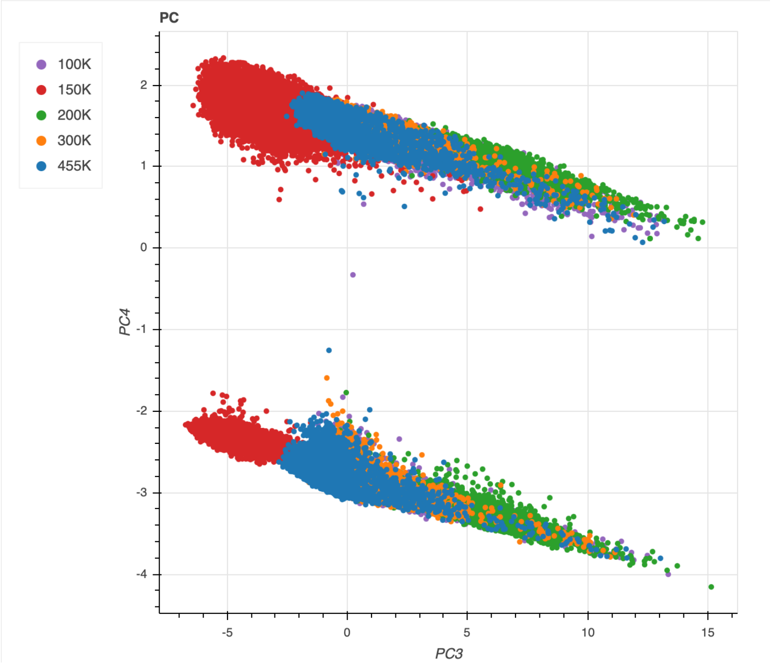


Fig. S5 | Platform inference using missingness PCA. PC3 vs PC4 colored by batch. Note that the batch names indicate the additional samples added from that batch. Thus, ‘100K’ refers to data tranche 1, ‘150K’ refers to samples added in tranche 1.5 (the first 50K samples released to the public), ‘200K’ refers to samples added in tranche 2, ‘300K’ refers to samples added in tranche 3, and '455K' refers to samples added in tranche 4. The separation in PC4 is driven by a common copy number variant.

*Relatedness inference*

We used Hail's *pc_relate* method to infer relatedness on SNVs that are autosomal, bi-allelic, common (allele frequency > 0.1%), high call rate (> 99%), and LD-pruned with a cutoff of $r^2 = 0.1$. We then used the *maximal_independent_set* method in Hail to keep the largest set of unrelated samples (samples without second degree or closer relationships; kinship > 0.1), prioritizing samples with greater mean depth. The related samples that were not in the maximal independent set were flagged.

The majority of the samples removed during sample QC were samples inferred to have a second-degree or closer relationship with other samples in the dataset. This finding is not unexpected, as previous studies have shown that the UK Biobank data contains a large number (approximately 30% at third-degree or greater) of related samples (Bycroft et al., 2018).

*Ancestry imputation*

We used a hybrid method to infer ancestry for sample QC. We projected the UK Biobank data onto the gnomAD population principal components (PCs) and then used a random forest classifier trained on gnomAD ancestry labels to assign ancestry to the UK Biobank samples. We observed that many samples labeled as African using this method were flagged as outliers by our downstream population-stratified outlier detection method. This seemed to be due to the fact that one cluster of samples labeled as African appeared highly admixed. To account for this, we ran a PCA on the UK Biobank samples and applied a clustering method (HDBSCAN). We found that this clustering method split the African labeled samples into additional clusters and reduced the number of samples flagged as outliers while also recapturing most of the same global population clusters observed in gnomAD.

As a result, we chose to assign ancestry using a hybrid of the projection onto gnomAD PCs and the UK Biobank specific PCA clustering: for any sample that was assigned to a cluster using the UK Biobank PCA, the sample was given that cluster as their ancestry assignment to preserve the sub-structure observed using clustering. Any sample that was not assigned to a cluster was given the label from the initial (gnomAD) PCA projection and random forest classification.

*Outlier detection*

We flagged any sample falling outside 4 median absolute deviations (MADs) from the median of any of the following metrics (stratified by population and tranche as a proxy for platform), which were calculated using hail's *sample_qc* method:

- Number of deletions
- Number of insertions
- Number of SNVs
- Insertion : deletion ratio
- Transition : transversion (TiTv) ratio
- Heterozygous : homozygous ratio

The final counts of samples is shown in Table S1.

Table S1 | Final sample counts passing QC. "nfe" refers to samples inferred as having non-Finnish European ancestry. Note that relatedness was run after hard filtering, so the total number of related and unrelated individuals is equal to the total number of samples less 683.

| Category | Related | Unrelated | All |
|---|---|---|---|
| Total | 29900 | 424114 | 454014 |
| Hard filtered | | | 683 |
| Outlier filtered | 184 | 2877 | 3061 |
| High quality | 29716 | 421237 | 450953 |
| High quality (nfe) | 28630 | 397740 | 426370 |

#### Variant QC

Variant filtering consisted of a combination of a random forest (RF) classifier and hard filters. We used the following training sets as true positives for training the random forest model:

- Omni - SNVs present on the Omni 2.5 genotyping array and found in 1000 Genomes data
- Mills - Indels present in the Mills and Devine data (Mills et al., 2006)
- Transmitted singletons - Variants found in only two individuals, which were a parent-offspring pair
- Sibling singletons - Variants found in only two individuals, which were a sibling pair
- Common (AF > 0.1%) and concordant (>90% non-reference concordance) array variants

For the false positive training set in the random forest model, we used variants that fail traditional GATK hard filters: QUAL by depth (QD) < 2, strand bias estimated using Fisher's exact test (FS) > 60, or root mean square mapping quality (MQ) < 30. We balanced the number of variants in the true positive and false positive training sets by randomly downsampling the false positive training set to the same number of variants found in the true positive training set. RF training was performed on only variants that fall within intervals that pass interval QC (described above; intervals where >85% of samples have a mean coverage >20X).

We used the following allele and site annotations as features in the random forest model (RF feature importance shown in Fig. S6):

- Allele type - SNV, indel
- Number of alleles - Total number of alleles present at the site
- Variant type - SNV, indel, multi-SNV, multi-indel, mixed
- Mixed site - Whether more than one allele type is present at the site
- Spanning deletion - Whether there is a spanning deletion (STAR_AC > 0) that overlaps the site
- Quality by depth - Sum of the non-reference genotype likelihoods divided by the sum of the depth in all carriers of that allele

- Read position RankSum - Rank Sum Test for relative positioning of reference versus alternate alleles within reads
- Mapping quality RankSum - Rank Sum Test for mapping qualities of reference versus alternate reads
- Strand bias odds ratio - Symmetric Odds Ratio test of 2x2 contingency table to detect strand bias
- Max probability of allele balance - Highest p-value for sampling the observed allele balance under a binomial model with p=0.5 (maximum across heterozygotes)

RF probability cutoffs for calling a variant PASS were chosen to maximize sensitivity and specificity based on criteria such as the number of de novo mutations found in the 224 trios in the dataset and precision-recall curves (Fig. S6B) in two truth samples present in our data (NA12878 and a pseudo-diploid sample (syndip); syndip was sequenced at Broad, not with the UK Biobank cohort). Final thresholds were RF true positive probability of 0.061 (approximately 90% of SNVs in well-covered intervals) for single nucleotide variants and 0.064 (approximately 75% of indels in well-covered intervals) for indels. Finally, we also excluded variants with two hard filters:

- Excess heterozygotes defined by an inbreeding coefficient < -0.3
- Variants where no sample had a high quality genotype (see Genotype QC below)

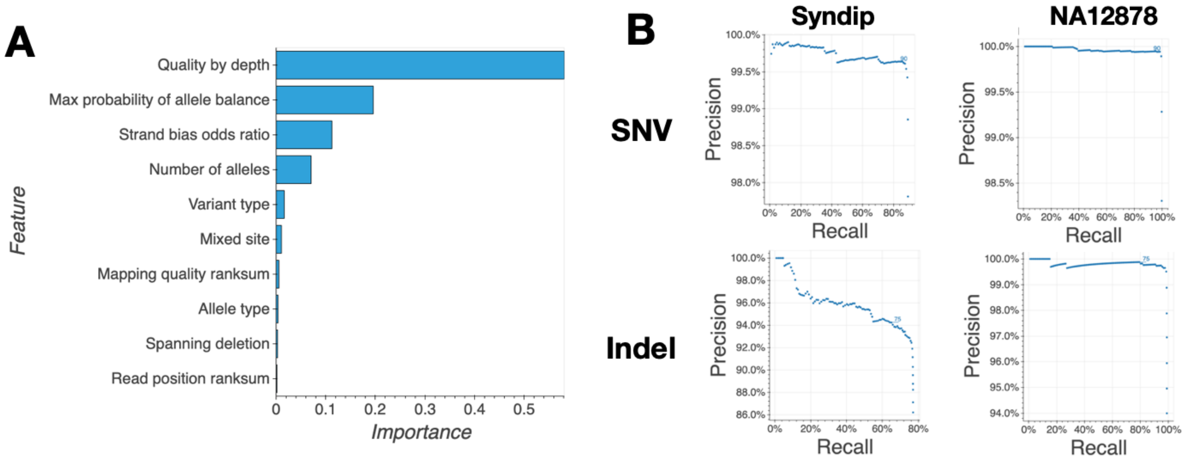


Fig. S6 | Variant QC (**A**): A summary of the features used in the random forests model and their relative importance in the model generated. (**B**): Precision and recall curves for the random forest classifier using two truth samples present in our data (NA12878 and syndip). The highlighted points at 90 for SNVs and 75 for indels indicate the cutoffs used for variant filtering.

**Genotype QC**

We filtered genotypes based on the previously defined “adj” criteria, with a modification for haploid calls on chrX and chrY for XY individuals. Specifically, we filtered to genotypes where depth >= 10 (5 for haploid calls), genotype quality >= 20, and minor allele balance > 0.2 for all alternate alleles for heterozygous genotypes.

**Annotations**

Variants were annotated using VEP v95 as implemented in Hail using the default parameters for GRCh38 (including LOFTEE (Karczewski et al., 2020)).

For downstream analyses, variants were grouped by Ensembl Gene ID and functional impact as follows:

- pLoF: High-confidence LoF variants (as indicated by LOFTEE), including stop-gained, essential splice, and frameshift variants, filtered according to a set of first principles as described at https://github.com/konradjk/loftee.
- missense|LC: Missense variants are grouped with in-frame insertions and deletions, as well as low-confidence LoF variants (filtered out by LOFTEE). The latter have a

frequency spectrum consistent with missense variation (Karczewski et al., 2020), and affect a set of amino acids in a similar fashion (e.g. a frameshift in the final exon).

- synonymous: All synonymous variants in the gene.

### Scaling association testing using Hail Batch

Jacqueline I. Goldstein, Daniel King, Konrad J. Karczewski, Benjamin M. Neale, Cotton Seed

To perform the association analysis using SAIGE-GENE, we developed a new scientific compute scheduler, Hail Batch. Hail Batch is a cloud-based, serverless, multi-tenant platform as a service (PaaS). To use Hail Batch, users construct a computational graph of jobs to be executed, called a batch, using a Python client library (or manually). The batch is then submitted to Hail Batch via a REST API. The Hail Batch scheduler both manages pools of worker virtual machines (VMs) on which to schedule user jobs and schedules jobs on those workers. Hail Batch includes a Web UI for monitoring batches and viewing individual job logs. The documentation can be found here: https://hail.is/docs/batch/index.html. Hail Batch is fully open source and is contained in the Hail project monorepo which can be found here: https://github.com/hail-is/hail/tree/main/batch.

**Motivation**

We built Hail Batch because we wanted a serverless solution with zero operational overhead for users. Based on our experience building Hail Query on Apache Spark, even with cloud platform managed services for running Spark like Google Dataproc, configuring, provisioning and managing compute clusters is a significant operational burden each user must bear (and scales with the number of users). In addition, by operating a multi-tenant compute cluster, we increase the utilization of resources amongst all users. The benefits are especially pronounced when users are working on iterative analyses (as opposed to batch processing) where they might need time to assess a result before moving on to subsequent analyses.

Bioinformatic tools come in a variety of forms, from standalone binaries and command line tools like GATK and SAIGE, to Python and R packages, to cloud-native analytics tools like Hail Query. We wanted a scheduling infrastructure that would support building data processing pipelines across all these tool modalities. Hail Batch currently supports executing containerized command line tools and serialized Python functions. A Hail Batch based backend for Hail Query which executes JVM bytecode is under development.

Finally, we wanted to support low scheduling overhead at a large scale so that users could decompose pipelines based on natural biological or data considerations rather than computational constraints. For example, running millions of relatively fast statistical tests (seconds or more) for permutation testing requires small scheduling overhead to be effective. The association analysis described here naturally decomposed into a job per megabase per phenotype, for a total of approximately 18 million jobs.

We follow the principle that the systems we build today should themselves become building blocks of the systems we build tomorrow just as the association pipelines described herein build on Hail Batch. Therefore, we wanted a system that had a native program programmatic interface so it could be used by other systems. For example, the Hail team's continuous integration (CI) system runs tests and deployments using Hail Batch. A future application of Hail Batch is serving as the underlying execution engine for an incremental joint caller service.

**User interface**

Hail Batch provides a Python library to construct and execute batches. A simple example is shown in Fig. S7. Rather than focus on the Python interface, which is described in detail in the documentation, we will focus on the main conceptual pieces of a Hail Batch computational graph.

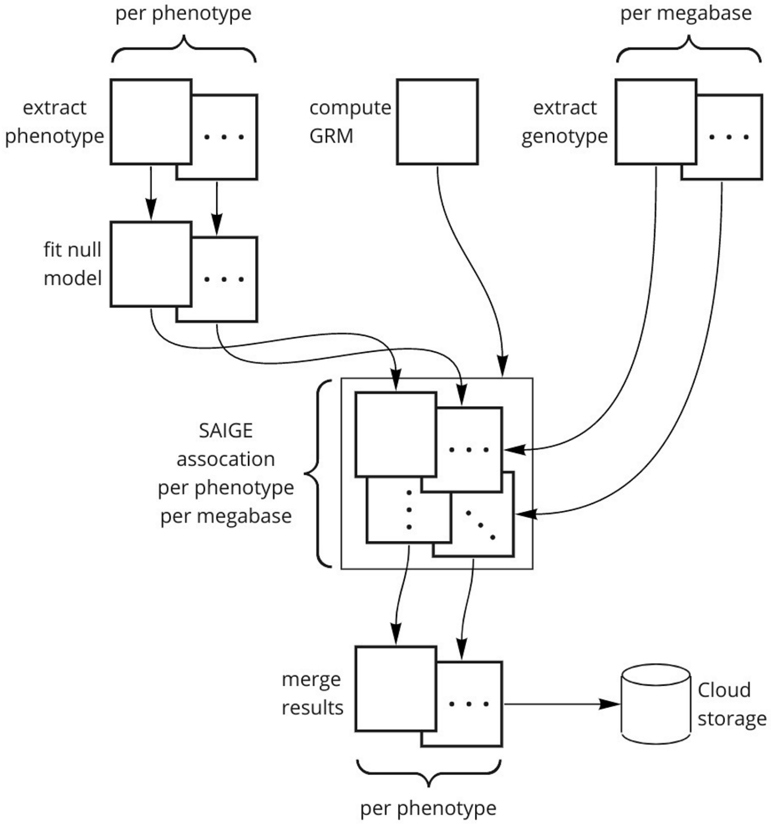


Fig. S7 | Hail Batch schematic for SAIGE association analysis. An example batch (the SAIGE pipeline used in this manuscript) is shown here.

A batch is the unit of submitted work. A batch primarily consists of (1) user-defined attributes, and (2) an ordered list of N jobs. The user-defined attributes are for searching and identifying batches. Jobs are the individual units of scheduling and can depend on previous jobs to form a directed acyclic graph (DAG) representing the full computational pipeline. The main parts of the job description are: (1) user-defined attributes, (2) dependencies, (3) inputs and outputs, (4) compute resources, and (5) executor configuration. Jobs do not run until all of their dependencies have completed. There are mechanisms for cancellation and controlling whether jobs run if their dependencies failed or were cancelled. Compute resources (CPU, memory, disk and machine type) describe resources needed to schedule the job, which will be provisioned by the autoscaler as needed. We refer the reader to the documentation for more details.

During the submission and execution of a batch, jobs pass through the following states: pending (dependencies are not complete), ready (dependencies are complete and the job is ready to be scheduled), creating (the job is provisioning resources for custom resource configurations), running (the job has been scheduled), and the terminal states: error (the executor failed to run the job), cancelled, failed (the job execution failed), or successful. The REST API and Web UI allow users to monitor running and historical batches and view individual job logs.

There are currently two backends for executing a batch: (1) a local backend that runs jobs locally and (2) a service backend that submits jobs to the Hail Batch service. Currently, the Hail Batch service supports the Google Cloud Platform.

**Implementation**

Hail Batch is written in Python and makes extensive use of asyncio. Its deployment consists of three parts: (1) the front-end **`batch`** service, (2) **`batch-driver`** service, and (3) a MySQL database instance. The **`batch`** service serves the Web UI and user-facing REST API queries. It is stateless and autoscales based on incoming traffic. The **`batch-driver`** runs the job scheduler, the autoscaler which provisions worker VMs, and the admin Web interface. Job and batch configuration state is stored in MySQL and job logs are stored in Google Storage. Hail Batch relies on a Hail PaaS **`auth`** service for session ID based authentication. The Hail PaaS services are deployed in Kubernetes and the autoscaler uses the Google Compute Engine (GCE) directly to provision workers. Hail Batch deployment is controlled by the **`ci`** (continuous integration) service. We also maintain Terraform scripts for bootstrap and disaster recovery of Hail PaaS installations.

The autoscaler in the driver service is organized in terms of instance pools. There are two types of instance pools, shared and private. There are three shared pools for the three main GCP machine types. Shared pools autoscale based on the pool size and the number of ready jobs that run in that pool. Batch pools can scale down to zero, but we also support a minimum size so small jobs can be dispatched immediately without waiting to spin up resources. Shared pool workers schedule jobs across all users and resource utilization is higher because instance startup and shutdown is generally amortized over many jobs. By default, preemptible n1-standard-16 machines are used, but jobs that use special machine types or require non-preemptible instances can run in a private pool which provisions a VM per job.

In addition to autoscaling, the Hail Batch scheduler implements a fair share algorithm across users to provide a responsive experience. Jobs are scheduled per user, in reverse order of the amount of resources already allocated. If there are not enough ready jobs to saturate a user's allocation, the unused allocation is made available to the remaining users. This means new jobs submitted by a user to an active cluster will scale up quickly to the user's share and all users enjoy a more responsive experience when the system is actively used.

We make a few remarks on the scheduler performance. The scheduler is able to schedule ~80 jobs/s. The maximum cluster size the scheduler can support is a function of the expected job length. The maximum cluster size is (average job length in second) * 80. Therefore, for the 15-30m SAIGE jobs used in the association analysis here, the scheduler is theoretically (and was practically) able to saturate a ~100K core pool of workers.

Hail Batch currently supports two executors on worker VMs: Docker and the Java Virtual Machine (JVM). The JVM executor is used by the Hail Query service. The Docker executor includes the details necessary to run a docker container: image, environment variables, command line, etc. Jobs run on the worker VMs in three steps: input, main and output. The input and output steps are responsible for copying between object storage and the local filesystem. For copying, we developed a pluggable Python asyncio filesystem abstraction and high-performance parallel copy management engine which supports local files, Google Cloud Storage, S3 and HTTP(S) (read-only). The main step executes the user's code.

### Association testing and quality control

Konrad J. Karczewski, Wenhan Lu, Jacqueline I. Goldstein, Daniel King, Wei Zhou, Cotton Seed, Benjamin M. Neale

##### Association testing framework

We performed association testing on the quality-controlled genotype data using SAIGE-GENE (Zhou et al., 2019), following the recommendations by the authors. We computed a genetic relatedness matrix (GRM) using a dataset sampled from allele frequency categories from the genotype MatrixTable considering only autosomal variants with a minimum call rate of 95%, including approximately 2,000 variants from each of Allele Count (AC) 1-5, AC 6-10, and AC 11-20; and approximately 10,000 variants from each of: AC 20-AF 0.1%, AF 0.1-1%, AF 1-10%, AF > 10%. These variants were LD-pruned to $r^2 = 0.1$, and exported into PLINK format. We computed a sparse GRM using step0 of SAIGE-GENE using the default parameters, with 2,000 markers used for the kinship matrix, and a relatedness cutoff of 0.125. We further created a “gene map” file for each megabase, which included information about the variants to be analyzed together in each group test. We included 3 groups: pLoF variants, including only those annotated as high-confidence by LOFTEE; missense-like variants, including missense variants and variants annotated as low-confidence by LOFTEE; and synonymous variants. The script for pre-processing is available at https://github.com/Nealelab/ukb_exomes/blob/master/hail/pre_process_saige_data.py.

The remainder of the process was parallelized using Hail Batch (Fig. S7). For each megabase of the genome, we exported a BGEN from the genotype MatrixTable with all variants that lie in genes that have a starting coordinate within that megabase. For each phenotype, we exported a flat file from the phenotype MatrixTable with the covariates used for analysis: age, sex, $age^2$, and 20 principal components, as well as interaction terms of age * sex and $age^2$ * sex. The phenotype data were combined with the sparse GRM computed above to fit a null model (without genotypes) using step1 of SAIGE-GENE with the default parameters and the covariates described above. Finally, we ran the genotype tests using the BGEN from each megabase, and the null model from each phenotype using step2 of SAIGE-GENE with the default parameters, plus maxMAFforGroupTest=0.01, maxMAF=0.5, LOCO=FALSE, and IsSingleVarinGroupTest=TRUE. The results across all megabases were loaded into two Hail Tables for each phenotype, for the group tests and single variant tests. The pipeline is available at https://github.com/Nealelab/ukb_exomes/blob/master/saige_exomes.py with helper scripts that can be found at https://github.com/Nealelab/ukb_common/blob/master/utils/saige_pipeline.py.

We combined the phenotype-level Hail Tables into a Hail MatrixTable using a hierarchical merge, along with phenotype metadata from SAIGE, resulting in one MatrixTable for the group tests and one for the single variant tests. We computed lambda GC values for each phenotype and gene (see below) using the *hl.methods.statgen._lambda_gc_agg* aggregator in Hail. These datasets are publicly released and serve the browser framework described below.

**Random phenotype analysis**

To test the asymptotic properties of our tests, we simulated 314 random normally distributed phenotypes in the 300K callset of this resource with a range of heritabilities using the sparseMVN package in R (Braun, 2015), using the genetic relatedness matrix generated by SAIGE. From these normal distributions, we simulated continuous phenotypes, as well as binary phenotypes with varying prevalences from $10^{-4}$ to 50%, with heritability of 1. We further generated a series of phenotypes of varying heritabilities (0.1, 0.2, 0.5) by introducing an additional noise component (rnorm) and weighting by the square root of the desired heritability. We performed association testing on these phenotypes and computed lambda GC values as above, and here, we show the qq-plots for the single-variant and group (SKAT-O and Burden) tests (Fig. S8).

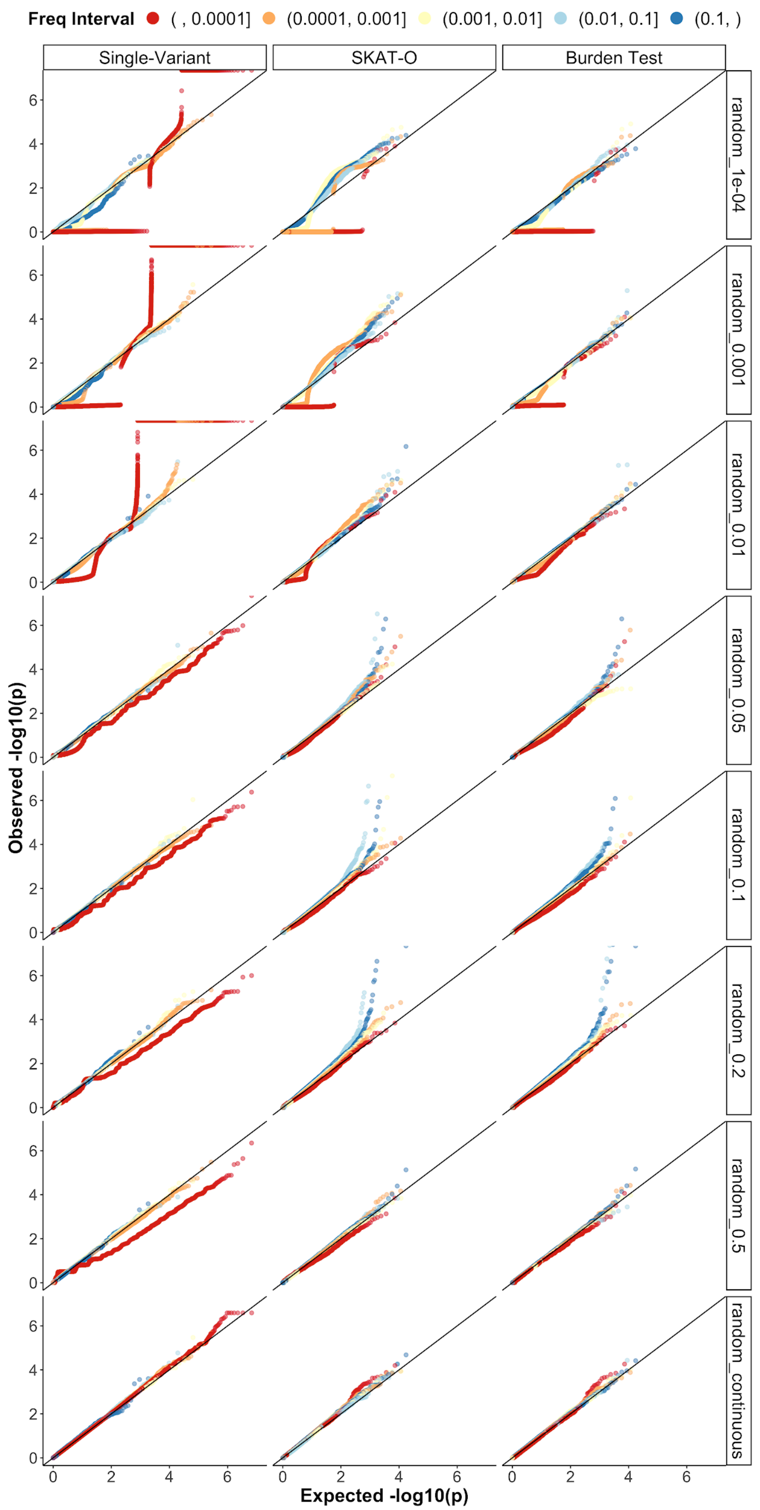


Fig. S8 | QQ-plots of randomly generated heritable (heritability = 100%) phenotypes for single-variant tests (left) and for group tests (SKAT-O, middle; and burden tests, right). The increasing prevalence of each binary phenotype is indicated by the label on the right (1e-4 to 0.5), followed by continuous traits.

In order to visualize the behavior of the tests across frequency strata in the association data, we compute the cumulative allele frequency (CAF) for each gene by aggregating the allele frequencies of variants of the same annotation group within each gene. We summarize the lambda GC metric for each trait for variants with a CAF category, which is shown in fig. S9. Notably, in Figs. S8-9, we can see increased instability of the QQ-plot and lambda values for rarer variants especially for rarer outcomes, suggesting the need for an allele frequency threshold for large-scale analyses. This is consistent with the minimum frequency and prevalence required to achieve statistical significance at this sample size (Fig. S10).

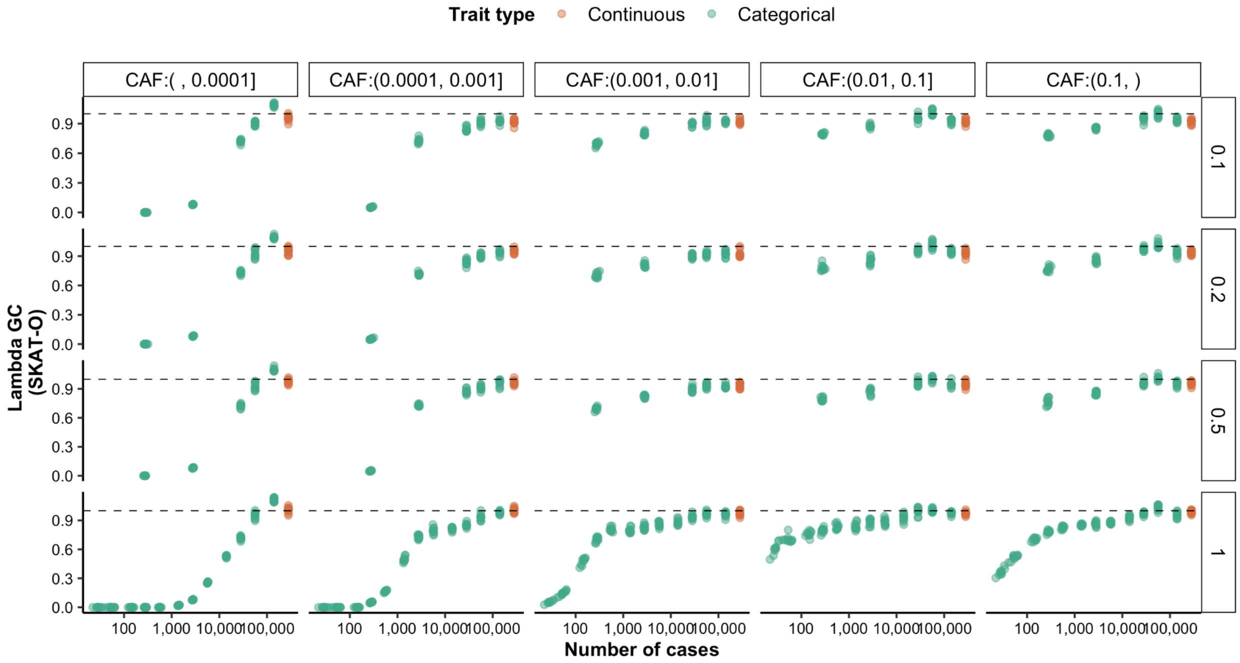


Fig. S9 | Lambda GC by cumulative allele frequency (CAF) by heritability. The heritability of the phenotypes are shown by the label on the right.

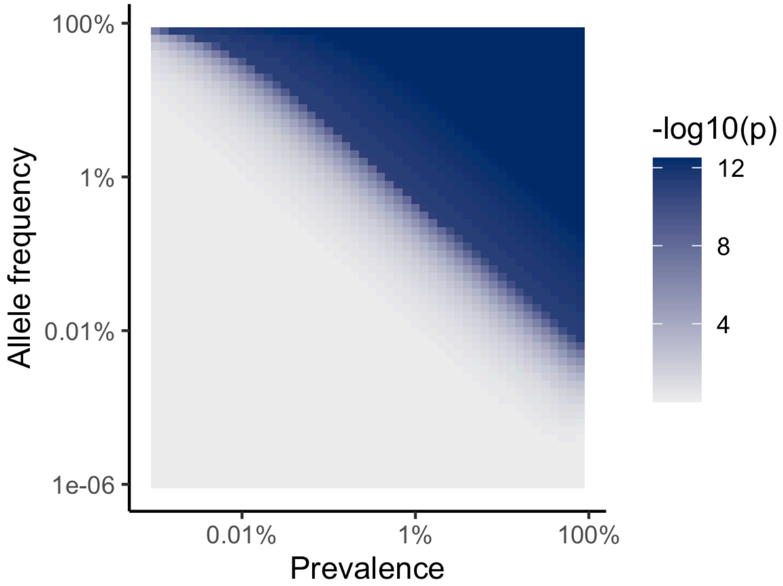


Fig. S10 | Power for rare variant associations. The minimum p-value possible from a protective mechanism with an odds ratio = 0: here, we compute the p-value of a chi-squared test of the case where the variant is absent from cases, while controls have a frequency as plotted. For the color-scale, a second logarithm is applied to p values below $10^{-10}$.

To compute an effective number of tests and thus a p-value threshold for each phenotype, we performed association testing between the genotype data and each of these phenotypes. The most significant p-value for each of the simulated continuous phenotypes is an estimator for the inverse of the effective number of tests: the median of this value across all simulated continuous phenotypes was $5 \times 10^{-6}$ for SKAT-O tests, $1.3 \times 10^{-5}$ for burden tests, and $1.6 \times 10^{-7}$ for single-variant tests. Thus, for downstream analyses, we computed the experiment-wise significance threshold as 0.05 * p, or: $2.5 \times 10^{-7}$ for SKAT-O tests, $6.7 \times 10^{-7}$ for burden tests, and $8 \times 10^{-9}$ for single-variant tests.

**Expected allele count filters**

Using the CAF for each gene and number of cases (n_cases) of the phenotypes, we then defined an element-level filter, the expected allele count, as CAF × n_cases (for continuous traits, this is the CAF × the number of individuals with a defined value). For each phenotype in each group test (SKAT-O, SKAT, and Burden test), we filtered the test results to genes from each of the expected AC intervals: [0, 5], (5, 50], (50, 500], (500, 5000], (5000, 50000], and (50000, ∞) and computed lambda GC values for each phenotype within each bin (Fig. S11). Lambda GC values for categorical phenotypes converge to around 1 as expected

AC increases Lambda GC values for continuous phenotypes are not significantly affected by the change of expected AC. A smaller number of cases and lower level of expected AC results in less stable values of lambda. Because of the highly deflated pattern of lambda GC values observed in genes with expected AC from 0 to 50, we filtered out summary statistics with expected AC < 50. For the variant-level results, we defined the expected AC as MAF × n_cases and computed lambda GC across different intervals. We observed a similar deflated pattern at low levels of expected AC and filtered out summary statistics with expected AC < 50.

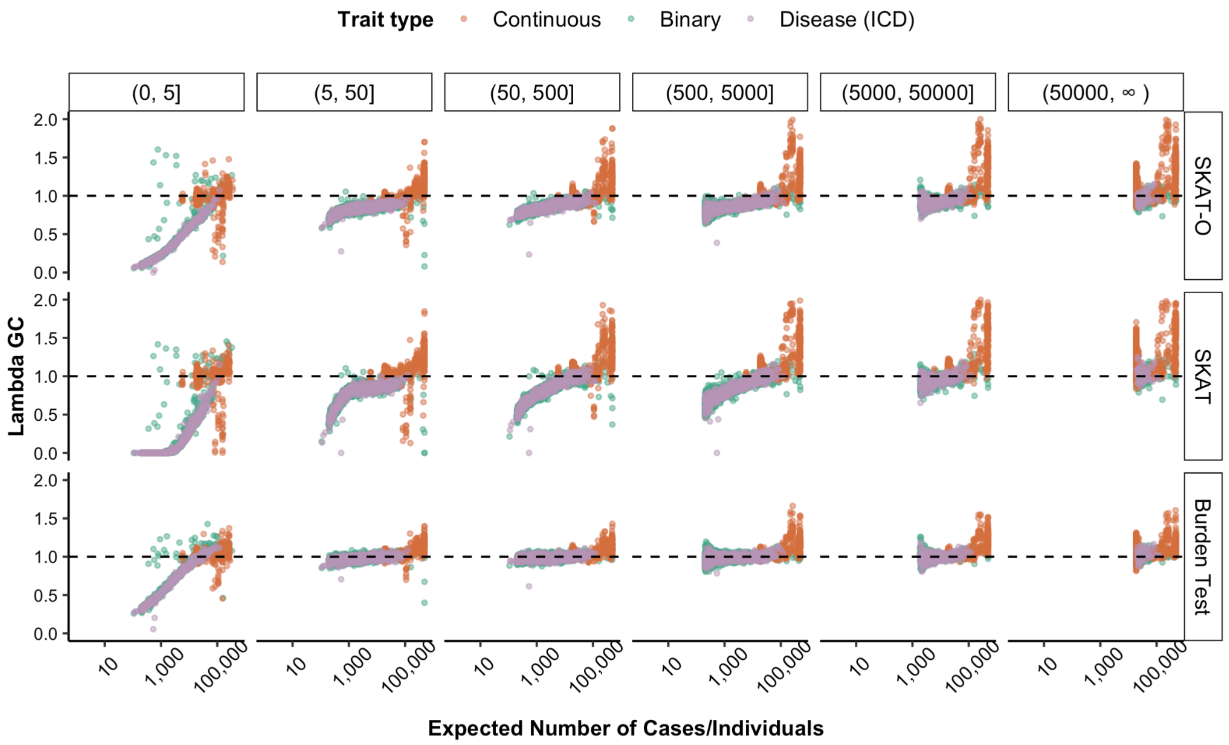


Fig. S11 | Lambda GC for each phenotype vs case count, split by expected AC interval for SKAT-O, SKAT and burden tests.

**Test calibration filtering**

Removing summary statistics with expected AC < 50 results in improved lambda GC values (fig. S12). Further, we filtered association statistics with standard errors (SE) of 0, as these were the result of a bug in the version of SAIGE-GENE used herein (which has since been corrected).

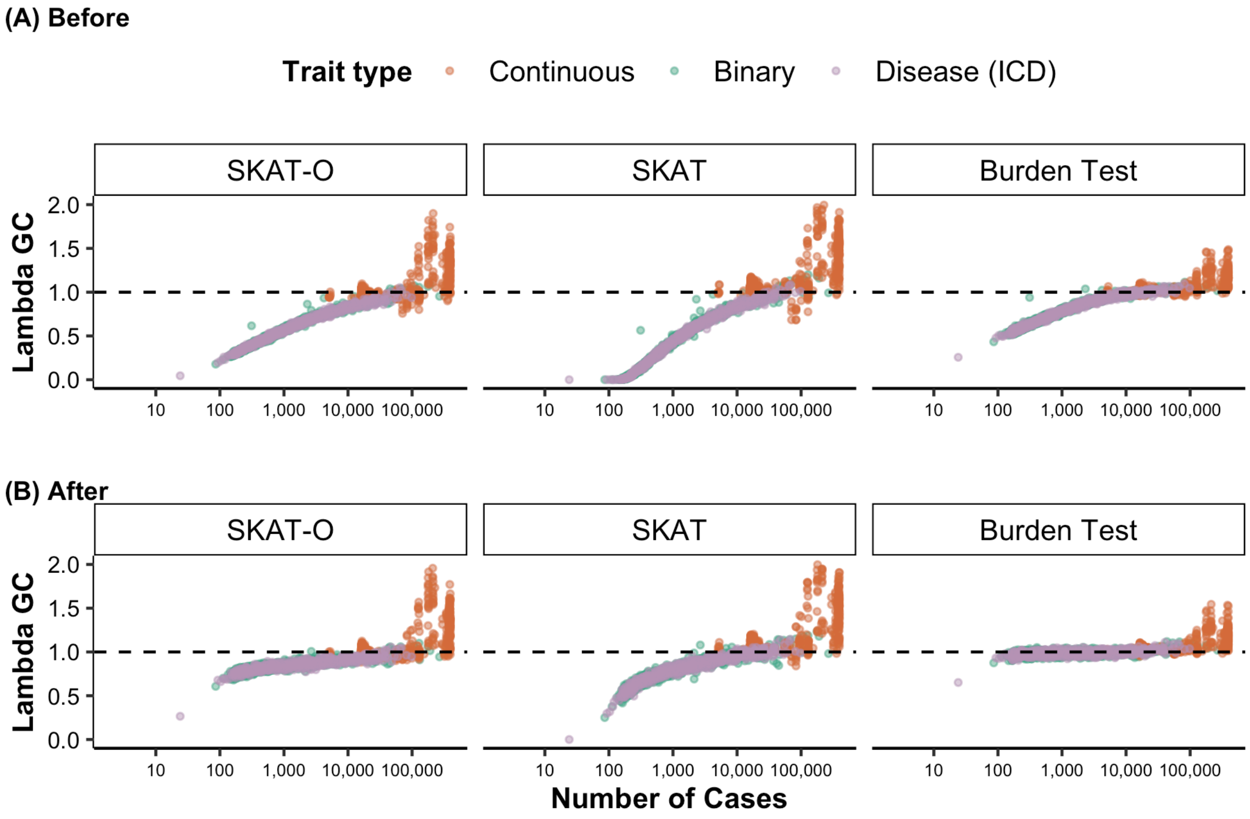


Fig. S12 | Lambda GC for each phenotype vs case count for SKAT-O, SKAT and burden tests, before and after filtering out summary statistics with expected AC < 50, number of variants tested < 2, and coverage < 20.

Finally, due to the large number of phenotypes available, we devised a metric for the calibration of individual genes, a lambda GC for each gene across phenotypes. For most genes, this metric is well-behaved for synonymous variants (95% range). However, it also marks outliers for removal, and appears to be correlated with mean sequencing coverage (Fig. S13).

Thus, we removed genes with coverage < 20 as well as genes that have a synonymous lambda GC < 0.75.

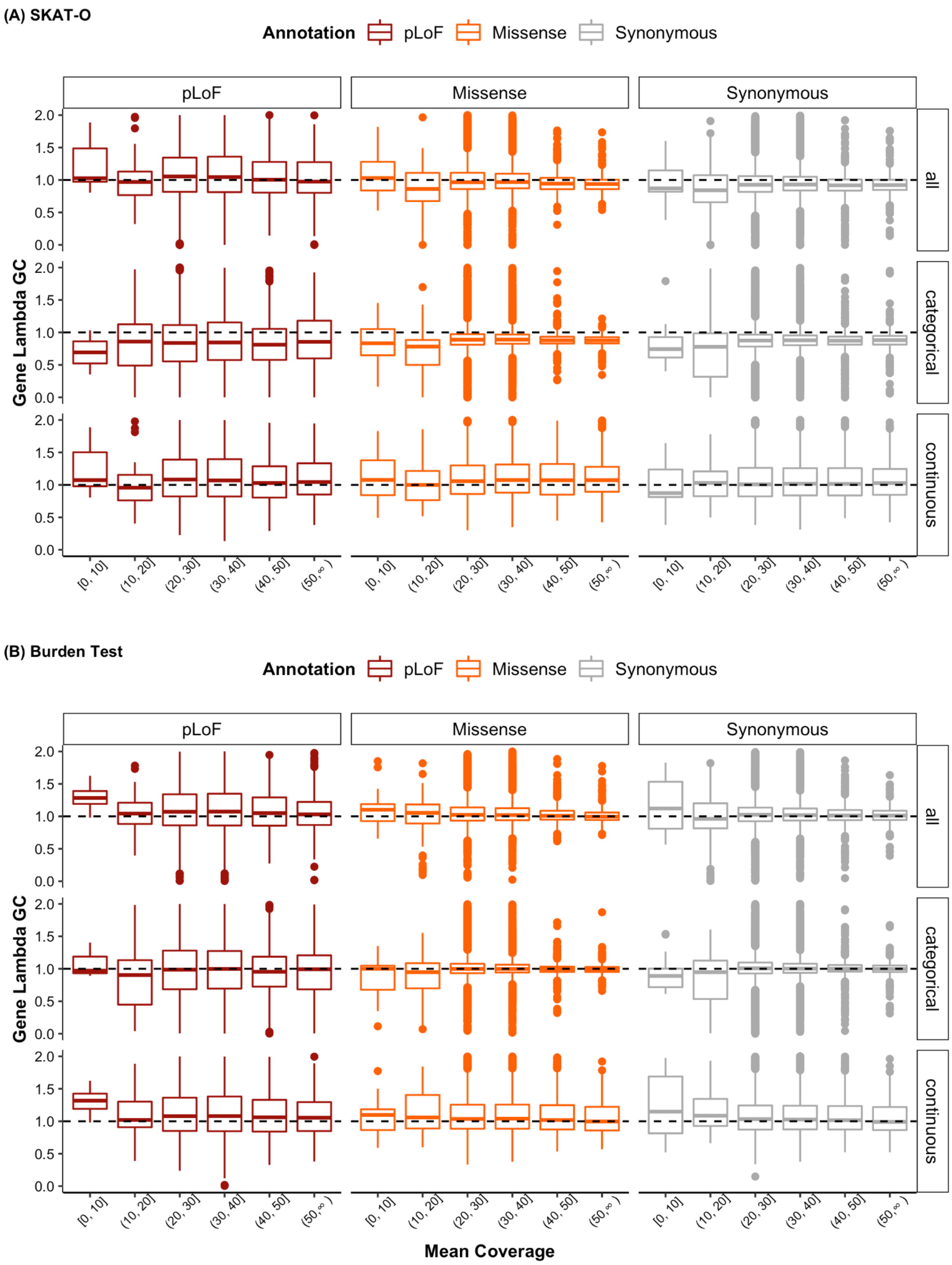

Fig. S13 | Coverage vs gene-based lambda GC, for SKAT-O (**A**) and burden tests (**B**).

After applying the above filters, we recompute lambda GC for each phenotype (Fig. S14) and gene (Fig. S15). For downstream analyses, we filter to phenotypes with lambda GC at least 0.75.

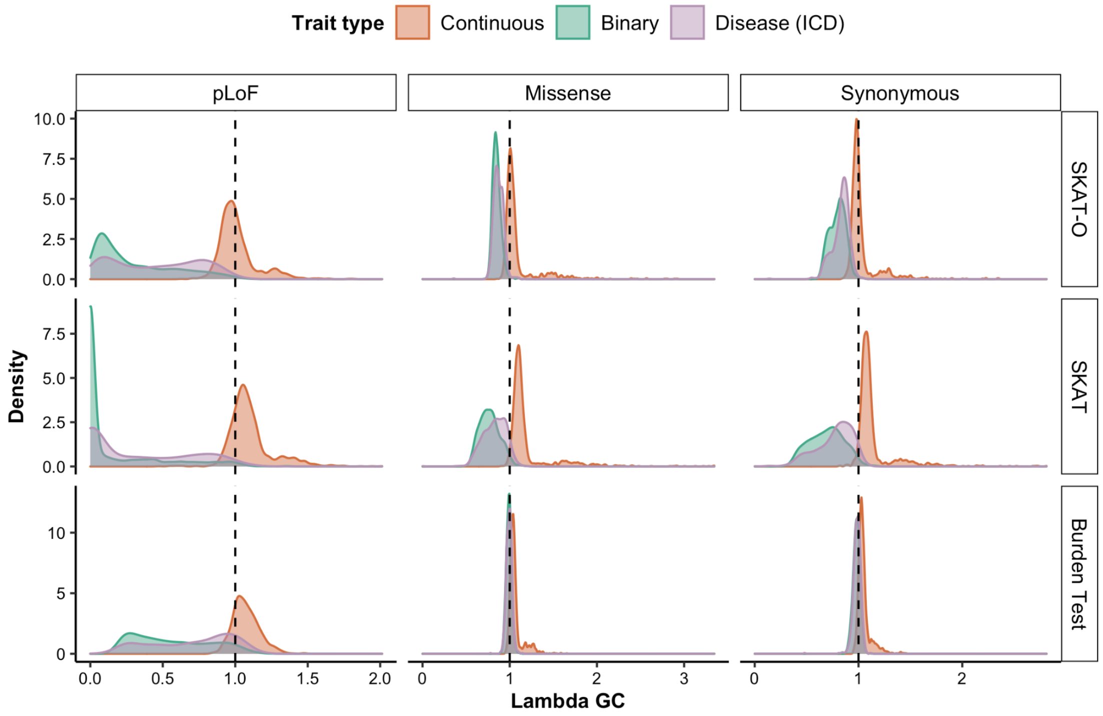


Fig. S14 | Lambda GC for each phenotype. A density plot of the distribution of lambda GC values for each phenotype is shown, broken down by trait type, test type, and set of variants used in the lambda calculation.

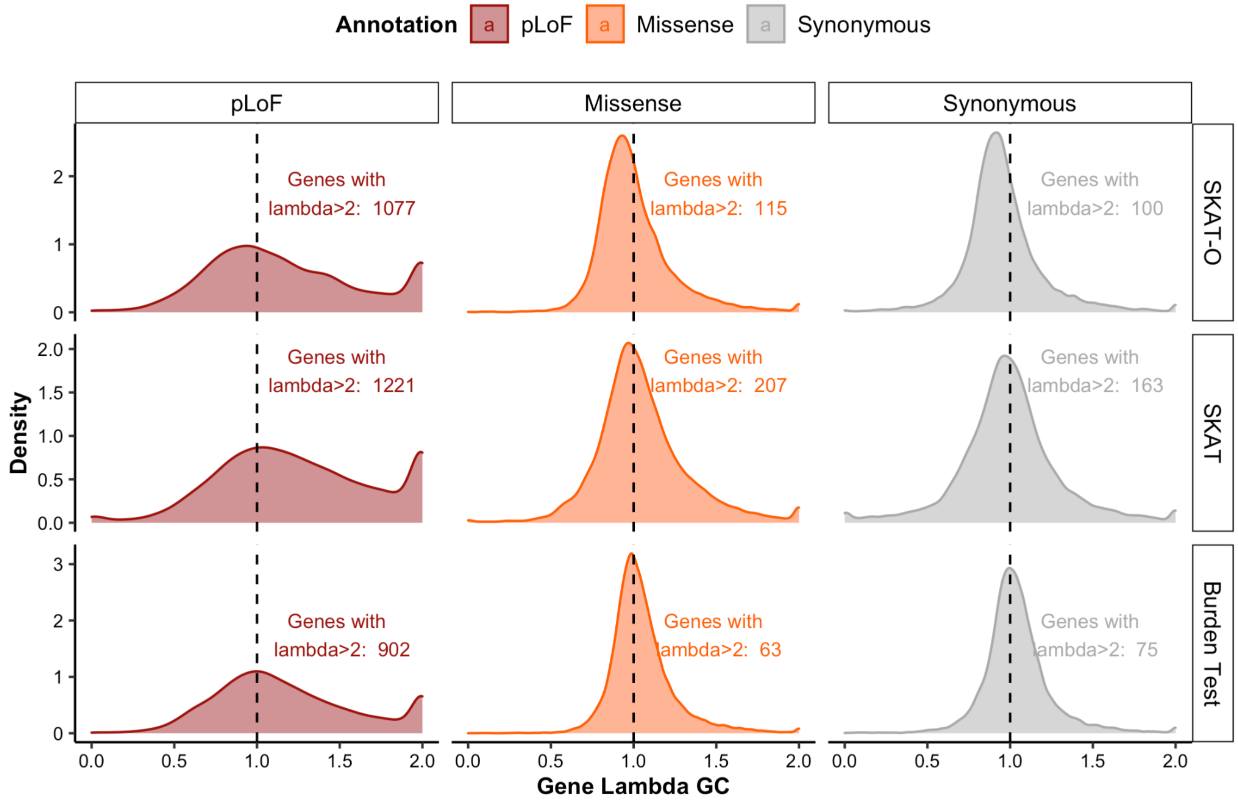


Fig. S15 | Lambda GC for each gene. A density plot of the distribution of lambda GC values for each gene is shown, broken down by test type and set of variants used in the lambda calculation.

#### Independent phenotypes

For large-scale analyses, we pruned to a set of relatively uncorrelated phenotypes. Using the UKB phenotype MatrixTable, we generated a pairwise correlation table using a matrix multiplication of the table and its transpose (Fig. S16A), and filtered the table to phenotype pairs with correlations ($r^2$) over 0.5. We then applied the *maximal_independent_set* function in Hail to the remaining phenotype pairs with a tie-breaker function preferring phenotypes with more cases, resulting in a set of 640 related phenotypes to remove from the dataset (Fig. S16B). A summary of the final QC steps is shown in Table S2.

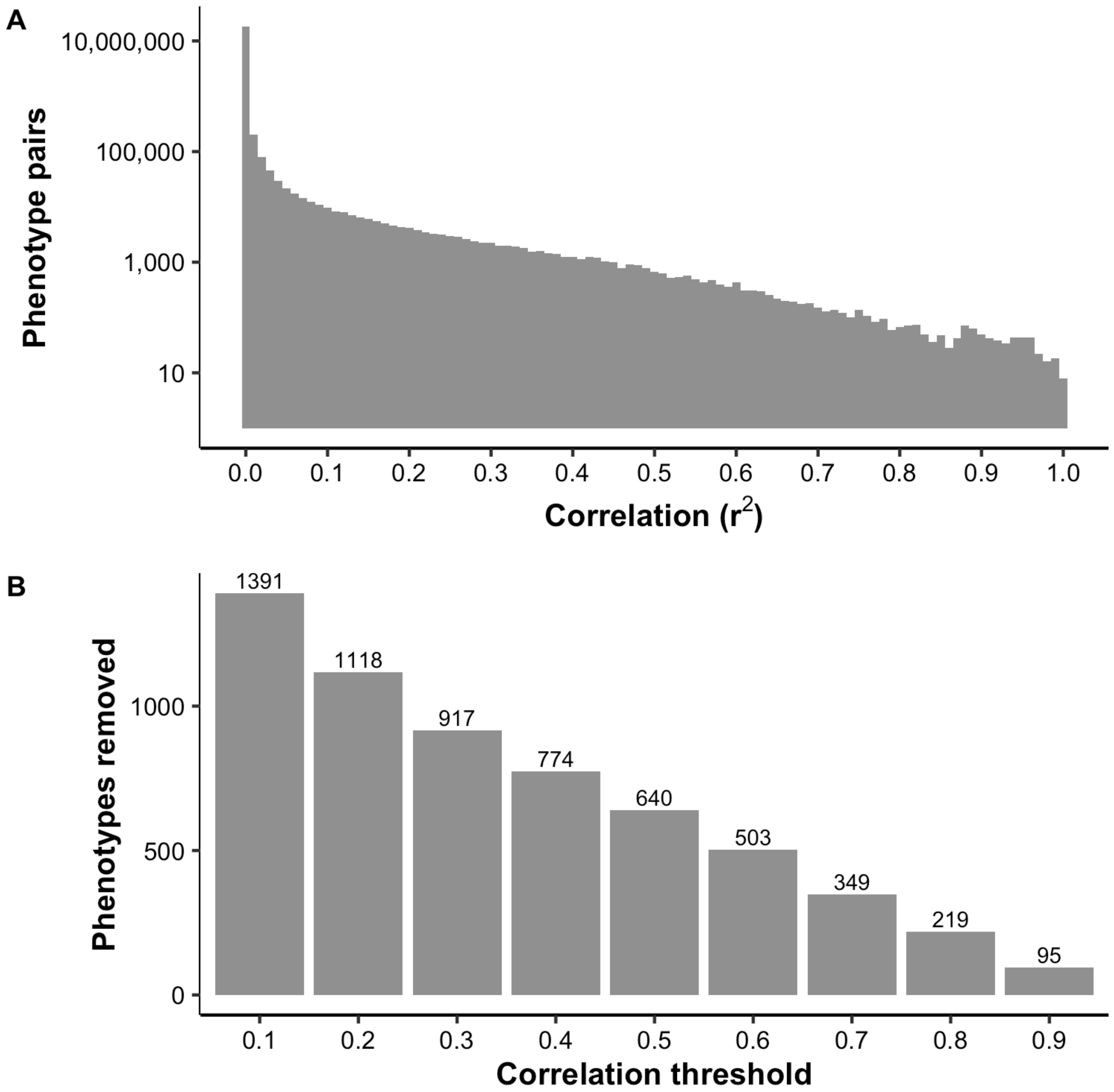


Fig. S16 | Independence of phenotypes. (**A**): A histogram of the number of phenotype pairs by correlation ($r^2$). (**B**): The number of phenotypes that would be removed by the maximum independent set method, by $r^2$ threshold.

Table S2 | QC of summary statistics. All filters are applied sequentially.

| | Description | Count (% Percentage Remaining) | | | |
|---|---|---|---|---|---|
| | | pLoF | Missense | Synonymous | Total |
| **Group (SKAT-O)** | Before filtering | 18,358 | 19,403 | 19,372 | 75,767 (Oth:18,634) |
| | Number of variants >= 2 | 17,876 (97.4%) | 19,392 (99.9%) | 19,355 (99.9%) | 75,251 (99.3%) (Oth:18,628) |
| | Mean coverage >= 20 | 17,370 (94.6%) | 18,791 (96.8%) | 18,768 (96.9%) | 72,999 (96.3%) (Oth:18,070) |
| | At least 1 phenotype with expected AC (CAF*n_cases) >= 50 | 10,790 (58.8%) | 18,741 (96.6%) | 18,660 (96.3%) | 66,221 (87.4%) (Oth:18,030) |
| | Lambda of the synonymous group > 0.75 | 9,636 (52.5%) | 16,320 (84.1%) | 16,354 (84.4%) | 58,056 (76.6%) (Oth:15,746) |
| **Variant** | Before filtering | 515,246 | 5,279,243 | 2,274,565 | 8,074,878 (NA: 5,824) |
| | Annotation defined | 515,246 | 5,279,243 | 2,274,565 | 8,069,054 (99.9%) |
| | At least 1 phenotype with expected AC (CAF*n_cases) >= 50 | 8,404 (1.6%) | 224,148 (4.2%) | 152,482 (6.7%) | 385,034 (4.8%) |
| | | **Continuous** | **Categorical** | **Disease (ICD)** | **Total** |
| **Phenotype (SKAT-O)** | Before filtering | 1,233 | 2,571 | 725 | 4,529 |
| | Lambda > 0.75 | 1,233 (100%) | 2,349 (91.4%) | 692 (95.4%) | 4,274 (94.4%) |
| | Correlation < 0.5 | 677 (54.9%) | 2,270 (88.3%) | 690 (95.2%) | 3,637 (80.3%) |

#### Comparison to known hits

We compared the significant associations with height discovered from our results with the 91 height-associated variants ($p < 2 \times 10^{-7}$) and 10 height-associated genes ($p < 5 \times 10^{-7}$) discovered in GIANT (Marouli et al., 2017). Among the 91 rare/low-frequency variants associated with adult height in GIANT, 50 of the variants were found to be associated with height at $p < 8 \times 10^{-9}$, and 78 variants were found to be associated with height at $p < 0.05$ in UK Biobank (Table S3-Table S4). Among the 10 genes associated with height in GIANT, FLNB (pLoF, missense|LC), NOX4 (missense|LC), OSGIN1 (missense|LC), and UGGT2 (pLoF) are found associated with height at $p < 2.5 \times 10^{-7}$ (SKAT-O) or $p < 6.7 \times 10^{-7}$ (Burden), and all genes were found at nominal significance ($p < 0.05$) for either missense or pLoF variants (Table S5).

Table S3 | Comparison to 32 rare (MAF < 1%) variants associated with adult height in GIANT; in UK Biobank, 21 of these variants are found to be associated with height at $p < 8 \times 10^{-9}$ (blue), and 29 are associated with height at $p < 0.05$ (light blue).

| Locus | Allele (Ref) | Allele (Alt) | Annotation | Gene | P-value | | | |
|---|---|---|---|---|---|---|---|---|
| | | | | | UK Biobank | GIANT | | |
| | | | | | | Discovery | Validation | Combined |
| 1:32673514 | G | C | missense | IQCC | 1.11E-10 | 7.92E-08 | 3.83E-06 | 1.34E-12 |
| 1:41540902 | G | A | missense | SCMH1 | 1.34E-27 | 1.58E-25 | 9.42E-13 | 1.35E-36 |
| 1:41618297 | G | A | missense | SCMH1 | 6.88E-24 | 1.92E-15 | 1.32E-08 | 1.80E-22 |
| 1:149902342 | C | T | missense | MTMR11 | 1.82E-14 | 4.16E-06 | 7.11E-06 | 3.03E-10 |
| 1:183495812 | A | G | missense | SMG7 | 5.47E-14 | 4.97E-11 | 8.94E-05 | 1.61E-14 |
| 1:223178026 | T | C | missense | DISP1 | NA | 1.11E-09 | 1.22E-06 | 1.27E-14 |
| 2:219920461 | T | A | missense | IHH | 1.17E-06 | 1.09E-15 | 1.48E-09 | 1.85E-23 |
| 2:220078652 | C | T | missense | ABCB6 | 3.13E-16 | 3.43E-13 | 4.40E-04 | 2.47E-15 |
| 3:46939587 | C | T | missense | PTH1R | 9.93E-09 | 1.30E-11 | 5.48E-10 | 1.14E-19 |

| | | | | | | | | |
|---|---|---|---|---|---|---|---|---|
| 4:73179445 | C | T | missense | ADAMTS3 | 5.40E-10 | 1.82E-08 | 1.32E-04 | 1.30E-11 |
| 4:120422407 | T | G | missense | PDE5A | 1.04E-10 | 7.50E-17 | 1.28E-08 | 2.65E-23 |
| 5:32784907 | G | A | missense | NPR3 | 3.93E-22 | 1.05E-08 | 1.78E-06 | 7.91E-14 |
| 5:64766798 | G | A | missense | ADAMTS6 | 9.39E-17 | 7.82E-09 | 1.37E-08 | 4.80E-16 |
| 5:127668685 | G | T | missense | FBN2 | 1.04E-30 | 2.47E-33 | 5.06E-20 | 1.47E-52 |
| 5:172755066 | C | A | missense | STC2 | 2.25E-34 | 5.69E-15 | 1.32E-17 | 1.15E-30 |
| 6:155450779 | A | G | missense | TIAM2 | NA | 1.45E-08 | 8.50E-01 | 3.96E-08 |
| 7:73482987 | G | A | missense | ELN | 1.48E-13 | 2.63E-06 | 1.51E-03 | 2.31E-08 |
| 8:135614553 | G | C | missense | ZFAT | 2.66E-45 | 4.42E-26 | 1.20E-14 | 6.12E-38 |
| 8:135622851 | G | A | missense | ZFAT | 4.76E-14 | 1.54E-12 | 5.94E-18 | 2.05E-28 |
| 11:27016360 | G | A | missense | FIBIN | 3.70E-08 | 5.79E-12 | 1.56E-03 | 3.26E-14 |
| 11:94533444 | G | A | missense | AMOTL1 | 1.96E-06 | 9.01E-16 | 3.84E-07 | 2.84E-21 |
| 12:58138971 | G | A | missense | TSPAN31 | 4.63E-01 | 8.26E-08 | 2.85E-03 | 5.50E-09 |
| 12:121756084 | G | A | missense | ANAPC5 | 4.32E-15 | 1.09E-11 | 1.44E-11 | 1.45E-21 |
| 15:44153571 | C | T | missense | WDR76 | 1.05E-04 | 1.56E-06 | 3.42E-04 | 2.32E-09 |
| 15:89424870 | G | T | missense | HAPLN3 | 1.51E-33 | 2.84E-13 | 2.43E-11 | 1.02E-22 |
| 16:31474091 | A | G | missense / splice acceptor | ARMC5 | 5.76E-10 | 5.88E-12 | 1.16E-03 | 1.62E-13 |
| 16:47684830 | C | A | missense | PHKB | 1.39E-06 | 3.96E-14 | 1.04E-01 | 3.43E-12 |
| 16:67470505 | G | A | missense | HSD11B2 | 4.15E-08 | 1.27E-07 | 3.38E-04 | 1.97E-10 |
| 16:84900645 | G | A | missense | CRISPLD2 | 5.32E-14 | 9.13E-12 | 4.34E-09 | 2.92E-19 |
| 16:84902472 | G | A | missense | CRISPLD2 | 2.66E-22 | 7.75E-14 | 3.49E-08 | 2.36E-20 |
| 16:88798919 | G | T | missense | PIEZO1 | 4.38E-17 | 5.27E-12 | 1.99E-08 | 8.68E-19 |
| X:66941751 | C | G | missense | AR | 1.06E-08 | 7.05E-07 | 7.12E-09 | 2.67E-14 |

Table S4 | Comparison to 59 low-frequency (MAF between 1% and 5%) variants associated with adult height in GIANT; in UK Biobank, 10 of the variants were not tested, 30 of these variants are found to be associated with height at $p < 8 \times 10^{-9}$ (blue), and 49 are associated with height at $p < 0.05$ (light blue).

| Locus | Allele (Ref) | Allele (Alt) | Annotation | Gene | P-value | | | |
|---|---|---|---|---|---|---|---|---|
| | | | | | UK Biobank | GIANT | | |
| | | | | | | Discovery | Validation | Combined |
| 1:51873967 | G | A | missense | EPS15 | 3.84E-18 | 5.07E-08 | 7.60E-11 | 2.56E-17 |
| 1:119427467 | A | C | missense | TBX15 | 6.10E-31 | 1.61E-24 | 4.19E-15 | 2.79E-36 |
| 1:150551327 | G | A | missense | MCL1 | 1.33E-25 | 2.16E-09 | 7.86E-12 | 1.55E-19 |
| 1:154987704 | C | T | missense | ZBTB7B | 1.82E-12 | 7.30E-17 | 4.46E-10 | 3.46E-25 |
| 1:180886140 | C | T | missense | KIAA1614 | 1.50E-05 | 1.41E-06 | 4.51E-04 | 2.63E-09 |
| 2:20205541 | C | T | missense | MATN3 | NA | 2.67E-23 | 6.60E-19 | 3.74E-41 |
| 2:219949184 | C | T | intron | NHEJ1 | NA | 5.96E-21 | 1.12E-15 | 8.20E-37 |
| 2:179474668 | G | A | missense | TTN | NA | 1.35E-07 | 2.15E-01 | 3.44E-07 |
| 2:233077064 | A | G | intron | DIS3L2 | NA | 2.35E-16 | 2.58E-15 | 6.46E-31 |
| 3:14214524 | G | A | missense | XPC | 1.22E-09 | 1.22E-08 | 1.68E-02 | 1.29E-08 |
| 3:47162886 | C | T | missense | SETD2 | 1.30E-08 | 2.24E-08 | 2.22E-07 | 1.65E-13 |
| 3:49162583 | C | T | missense | LAMB2 | 2.72E-37 | 3.28E-12 | 1.33E-16 | 3.49E-27 |
| 3:98600385 | T | C | missense | DCBLD2 | 4.69E-04 | 1.23E-07 | 5.62E-05 | 1.68E-12 |
| 4:5016883 | G | A | missense | CYTL1 | 8.93E-18 | 2.01E-17 | 6.68E-11 | 1.86E-25 |
| 4:87730980 | C | T | missense | PTPN13 | 6.71E-36 | 1.94E-19 | 1.38E-15 | 9.43E-32 |
| 4:135121721 | T | C | missense | PABPC4L | 4.83E-07 | 1.39E-13 | 1.33E-04 | 7.54E-16 |
| 4:144359490 | C | T | missense | GAB1 | 8.42E-07 | 1.04E-08 | 3.24E-04 | 4.29E-12 |

| | | | | | | | | |
|---|---|---|---|---|---|---|---|---|
| 4:154557616 | C | T | missense | TMEM131L | 4.32E-08 | 7.75E-08 | 5.75E-06 | 2.18E-12 |
| 5:102338811 | A | G | missense | PAM | NA | 3.76E-06 | 8.47E-06 | 1.63E-10 |
| 5:126250812 | C | T | missense | MARCH3 | 5.87E-05 | 4.25E-08 | 2.45E-03 | 1.67E-10 |
| 5:135288632 | A | G | missense | LECT2 | 7.90E-06 | 1.02E-07 | 4.77E-04 | 1.36E-09 |
| 5:172196752 | A | G | missense | DUSP1 | 6.30E-14 | 4.00E-14 | 1.26E-06 | 1.93E-20 |
| 5:176637471 | G | A | missense | NSD1 | 3.58E-23 | 2.38E-17 | 2.62E-12 | 4.27E-30 |
| 5:176722005 | G | A | missense | NSD1 | 1.01E-37 | 1.86E-26 | 8.42E-18 | 2.32E-41 |
| 6:30851933 | G | A | intron | DDRI | NA | 1.11E-08 | 1.24E-05 | 4.64E-13 |
| 6:34730395 | C | T | synonymous | SNRPC | 5.24E-52 | 9.21E-33 | 9.59E-31 | 3.45E-60 |
| 6:41903798 | C | A | missense | CCND3 | 1.74E-41 | 5.51E-17 | 3.41E-08 | 1.28E-22 |
| 7:99489571 | G | A | 3'UTR | TRIM4 | NA | 3.28E-10 | 2.26E-07 | 1.40E-17 |
| 7:100490077 | G | A | synonymous | ACHE | 3.34E-06 | 8.59E-10 | 2.92E-02 | 2.98E-10 |
| 7:135123060 | G | C | missense | CNOT4 | 4.59E-20 | 2.31E-17 | 5.04E-10 | 3.90E-26 |
| 8:42226805 | C | G | missense | POLB | 8.53E-05 | 1.95E-06 | 1.30E-02 | 1.88E-07 |
| 9:34660864 | C | T | missense | IL11RA | 7.28E-11 | 5.20E-13 | 4.42E-03 | 4.01E-13 |
| 9:95063947 | C | T | missense | NOL8 | 3.67E-04 | 2.56E-06 | 3.45E-02 | 3.33E-06 |
| 10:79580976 | G | A | missense | DLG5 | 1.68E-21 | 2.72E-11 | 5.15E-11 | 7.66E-20 |
| 10:97919011 | A | G | missense | ZNF518A | 1.29E-05 | 9.94E-08 | 3.05E-03 | 3.91E-09 |
| 11:65715204 | G | A | missense | TSGA10IP | 2.23E-41 | 1.82E-21 | 1.41E-23 | 1.52E-43 |
| 12:7548996 | C | G | missense | CD163L1 | 1.05E-03 | 4.11E-08 | 6.68E-02 | 1.87E-08 |
| 12:69140339 | G | C | missense | SLC35E3 | 1.87E-10 | 1.13E-09 | 5.UE-04 | 1.29E-11 |
| 12:104408832 | T | C | missense | GLT8D2 | NA | 8.72E-10 | 5.82E-10 | 1.60E-17 |
| 13:50842259 | G | A | intron | DLEU1 | NA | 2.33E-37 | 7.02E-25 | 5.66E-57 |

| | | | | | | | | |
|---|---|---|---|---|---|---|---|---|
| 14:23313633 | G | A | missense | MMP14 | 5.63E-08 | 1.72E-08 | 7.81E-09 | 3.27E-16 |
| 14:24707479 | G | A | missense | GMPR2 | 1.38E-16 | 3.67E-16 | 1.34E-11 | 2.13E-29 |
| 14:45403699 | C | A | missense | KLHL28 | 1.53E-07 | 1.55E-06 | 4.13E-04 | 3.05E-09 |
| 14:70633411 | C | T | missense | SLC8A3 | 4.05E-11 | 2.49E-11 | 2.02E-06 | 2.03E-16 |
| 14:94844947 | C | T | missense | SERPINA1 | 1.53E-100 | 1.39E-45 | 2.50E-34 | 1.72E-75 |
| 14:101349454 | G | T | missense | RTL1 | 7.09E-12 | 1.17E-11 | 2.12E-04 | 2.50E-15 |
| 15:34520687 | T | C | missense | EMC4 | 6.45E-02 | 1.16E-06 | 2.19E-02 | 1.60E-07 |
| 15:72462255 | C | T | missense | GRAMD2A | 2.04E-27 | 8.72E-17 | 3.66E-13 | 1.28E-27 |
| 15:89388905 | C | T | synonymous | ACAN | 1.61E-150 | 4.30E-72 | 1.08E-56 | 3.79E-130 |
| 16:4812705 | A | G | missense | ZNF500 | 4.21E-10 | 8.61E-17 | 2.34E-07 | 2.89E-21 |
| 16:24804954 | A | T | missense | TNRC6A | 3.87E-13 | 1.08E-09 | 1.65E-07 | 1.90E-15 |
| 16:67409180 | G | A | missense | LRRC36 | 2.22E-19 | 1.08E-18 | 3.91E-13 | 6.40E-31 |
| 17:67081278 | A | G | missense | ABCA6 | 5.70E-14 | 2.17E-06 | 5.58E-07 | 5.57E-12 |
| 18:74980601 | A | T | missense | GALR1 | 6.28E-07 | 3.60E-18 | 3.64E-05 | 5.11E-19 |
| 19:45296806 | C | T | missense | CBLC | 5.91E-03 | 1.48E-07 | 1.19E-02 | 2.96E-08 |
| 19:55879672 | C | T | missense | IL11 | 5.24E-47 | 1.02E-57 | 2.28E-23 | 5.32E-81 |
| 19:55993436 | G | T | missense | ZNF628 | 9.38E-47 | 2.28E-18 | 1.17E-18 | 6.33E-34 |
| 22:28501414 | C | T | missense | TTC28 | NA | 9.47E-11 | 3.24E-09 | 3.93E-19 |
| 22:42095658 | T | G | missense | MEI1 | 4.63E-04 | 2.25E-08 | 6.59E-03 | 3.70E-10 |

Table S5 | Comparison to 10 genes associated with adult height in GIANT. In UK Biobank, FLNB (pLoF, missense|LC), NOX4 (missense|LC), OSGIN1 (missense|LC), and UGGT2 (pLoF) reach our genome-wide significance threshold (SKAT-O $p < 2.5 \times 10^{-7}$ ; Burden $p < 6.7 \times 10^{-7}$) (blue), but all are found nominally significant for either pLoF or missense variants (light blue).

| Gene | UK Biobank | | | GIANT P-value | | | |
|---|---|---|---|---|---|---|---|
| | **Annotation** | **Burden Test** | **SKAT-O** | **SKAT-Broad** | **VT-Broad** | **SKAT-Strict** | **VT-Strict** |
| B4GALNT3 | missense\|LC | 5.49E-03 | 6.64E-03 | 2.40E-05 | 1.90E-05 | 1.80E-05 | **3.10E-07** |
| | pLoF | 3.25E-06 | 4.76E-06 | | | | |
| | synonymous | 6.46E-01 | 2.33E-01 | | | | |
| CCDC3 | missense\|LC | 3.80E-02 | 9.03E-03 | 6.30E-04 | 6.30E-06 | 3.00E-07 | **5.40E-09** |
| | pLoF | 7.55E-01 | 6.25E-01 | | | | |
| | synonymous | 4.00E-01 | 5.62E-01 | | | | |
| CRISPLD1 | missense\|LC | 8.61E-02 | 1.37E-01 | 2.20E-07 | **6.70E-11** | 8.50E-06 | 8.90E-07 |
| | pLoF | 5.00E-03 | 6.84E-03 | | | | |
| | synonymous | 3.81E-02 | 6.55E-02 | | | | |
| CSAD | missense\|LC | 3.57E-03 | 6.54E-03 | 2.30E-08 | **2.40E-09** | 0.83 | 0.59 |
| | pLoF | 3.33E-01 | 4.63E-01 | | | | |
| | synonymous | 8.84E-02 | 6.97E-04 | | | | |
| FLNB | missense\|LC | 2.99E-08 | 2.12E-08 | 2.20E-06 | 5.10E-04 | **2.40E-09** | 3.20E-06 |
| | pLoF | 5.51E-11 | 9.35E-11 | | | | |
| | synonymous | 7.37E-01 | 3.00E-02 | | | | |
| G6PC | missense\|LC | 5.77E-01 | 6.64E-02 | 1.30E-05 | **3.60E-08** | 5.50E-06 | 1.30E-06 |
| | pLoF | 3.03E-03 | 5.28E-03 | | | | |
| | synonymous | 4.82E-01 | 4.06E-01 | | | | |
| NOX4 | missense\|LC | 1.47E-10 | 5.27E-14 | 5.10E-06 | **1.40E-07** | NA | NA |
| | pLoF | 3.01E-04 | 5.04E-04 | | | | |
| | synonymous | 8.31E-01 | 2.39E-01 | | | | |
| OSGIN1 | missense\|LC | 8.14E-04 | 9.28E-11 | **4.30E-11** | 4.50E-05 | 0.19 | 0.18 |

| | | | | | | | |
|---|---|---|---|---|---|---|---|
| | pLoF | 7.40E-01 | 7.76E-02 | | | | |
| | synonymous | 4.65E-01 | 6.53E-01 | | | | |
| SNED1 | missense\|LC | 3.67E-02 | 6.24E-02 | 1.90E-05 | **4.30E-09** | NA | NA |
| | pLoF | NA | 3.69E-01 | | | | |
| | synonymous | 1.32E-01 | 2.16E-01 | | | | |
| UGGT2 | missense\|LC | 4.47E-05 | 1.09E-04 | 3.00E-05 | **2.60E-07** | 2.30E-05 | 4.80E-07 |
| | pLoF | 7.84E-09 | 6.51E-09 | | | | |
| | synonymous | 8.76E-02 | 1.48E-01 | | | | |

We compared the effect sizes from our single-variant test results to those of GIANT. Among 1,330 ExomeChip variants with p-value < 2 x $10^{-7}$ in the GIANT European-ancestry meta-analysis, 496 variants are tested in the UK Biobank data. The effect sizes of this group of shared variants are consistent between the two datasets, with a slight attenuation in results from UK Biobank, consistent with a degree of winner's curse (Fig. S17).

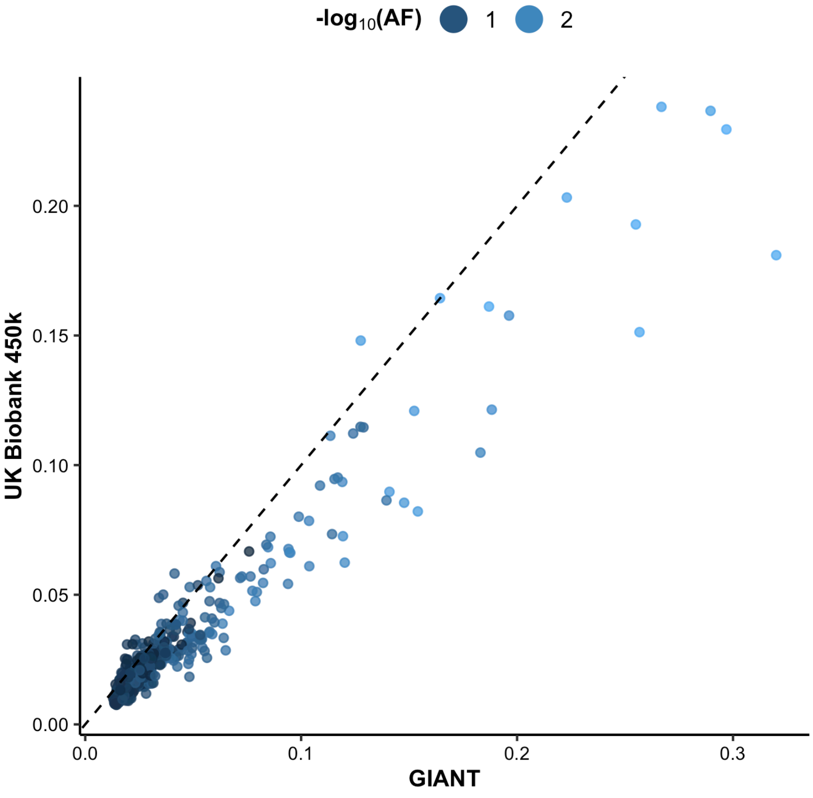

Fig. S17 | Comparison of effect sizes between UK Biobank and GIANT for height. The y = x line is shown for reference.

Finally, we compared associations for 7 red blood cell phenotypes discovered in our results with 20 associations ($p < 5 \times 10^{-9}$) between missense variants and red blood cell phenotypes discovered by TOPMed (Hu et al., 2021) and find that 19 out of 20 are replicated at $p < 0.05$ with 9 of these associated at $p < 8 \times 10^{-9}$ (Table S6).

Table S6 | Comparison of 20 associations between missense variants and 7 major red blood cell phenotypes discovered at the genome-wide significant loci of the marginal tests in TOPMed; in UK Biobank, 9 of these associations are significant at $p < 8 \times 10^{-9}$ (blue), and 19 are found significant at $p < 0.05$ (light blue).

| Phenotype | UKB phenocode | Locus | Allele (Ref) | Allele (Alt) | Gene | Annotation | P-value | |
|---|---|---|---|---|---|---|---|---|
| | | | | | | | TOPMed | UK Biobank |
| hematocrit (HCT) | 30030: hematocrit percentage | chr6:26092913 | G | A | HFE | missense | 6.40E-17 | 5.05E-174 |
| | | chr22:37066896 | A | G | TMPRSS6 | missense | 1.03E-26 | 3.28E-182 |
| | | chrX:154536002 | C | T | G6PD | missense | 3.36E-22 | 1.61E-03 |
| hemoglobin (HGB) | 30020: hemoglobin concentration | chr6:26092913 | G | A | HFE | missense | 2.16E-30 | 1.00E-300 |
| | | chr22:37066896 | A | G | TMPRSS6 | missense | 3.16E-51 | 1.00E-300 |
| | | chrX:154536002 | C | T | G6PD | missense | 1.47E-28 | 2.02E-02 |
| mean corpuscular hemoglobin (MCH) | 30050: Mean corpuscular hemoglobin | chr11:5227003 | C | T | HBB | missense | 1.24E-23 | 2.06E-02 |
| | | chrX:154536002 | C | T | G6PD | missense | 2.12E-48 | 7.91E-03 |
| mean corpuscular hemoglobin concentration (MCHC) | 30060: Mean corpuscular hemoglobin concentration | chr6:26092913 | G | A | HFE | missense | 9.52E-17 | 3.59E-246 |
| | | chr11:5227003 | C | T | HBB | missense | 4.29E-43 | 2.37E-02 |
| | | chr22:37066896 | A | G | TMPRSS6 | missense | 3.25E-26 | 5.70E-189 |
| | 30040: | chr1:247876149 | C | T | TRIM58 | missense | 1.77E-16 | 1.33E-118 |

| | | | | | | | | |
|---|---|---|---|---|---|---|---|---|
| mean corpuscular volume (MCV) | mean corpuscular volume | chr11:5227003 | C | T | HBB | missense | 1.36E-64 | 2.87E-04 |
| | | chr16:67184472 | T | C | EXOC3L1 | missense / synonymous | 2.13E-09 | 7.48E-29 |
| | | chrX:154536002 | C | T | G6PD | missense | 3.96E-82 | 5.87E-02 |
| red blood cell count (RBC) | 30010: red blood cell (erythrocyte) count | chr11:5227003 | C | T | HBB | missense | 2.44E-22 | 1.49E-02 |
| | | chrX:154536002 | C | T | G6PD | missense | 3.72E-82 | 1.27E-04 |
| red blood cell width (RDW) | 30070: red blood cell (erythrocyte) distribution width | chr6:26092913 | G | A | HFE | missense | 5.80E-15 | 1.00E-300 |
| | | chr11:5227003 | C | T | HBB | missense | 1.59E-10 | 1.51E-02 |
| | | chrX:154536002 | C | T | G6PD | missense | 8.27E-106 | 1.87E-04 |

### Analysis of summary statistics

Wenhan Lu, Ellen Tsai, Mark J. Daly, Benjamin M. Neale, Konrad J. Karczewski

We performed all downstream analyses in Hail using the single-variant and group-test MatrixTables as described above. For these analyses, we assess the proportion of genes or variants that reach our p-value threshold for at least one trait. Notably, when simply computing this metric across functional classes, seemingly paradoxically, we observe that the proportion of associations does not correlate with the expected deleteriousness of the variants (pLoF, followed by missense, followed by synonymous) for gene-based or single-variant tests (Fig. S18).

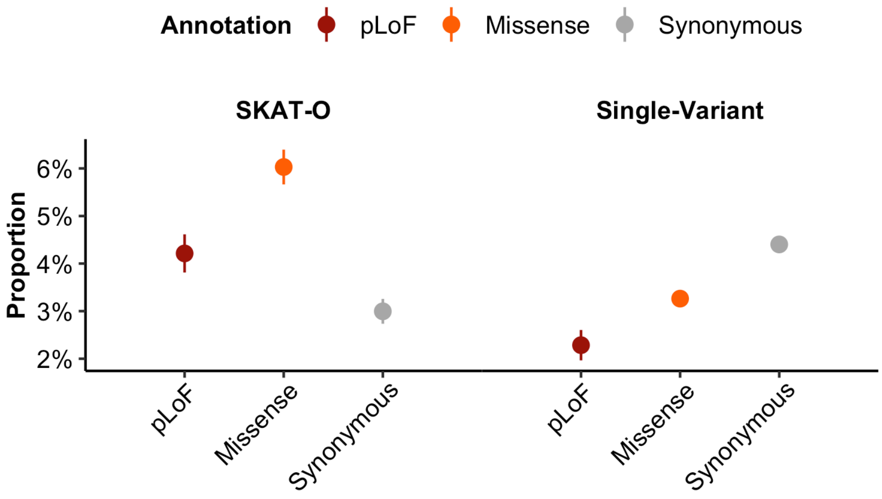


Fig. S18 | The proportion of genes (**A**) and variants (**B**) associated with at least one trait, broken down by functional class.

However, considering the observation that functional pLoF variants tend to be rare, and thus, have lower power for genetic discovery, we performed a series of analyses incorporating the frequency of variants and found that the former observation is due to Simpson's paradox. First, when separating genes by frequency strata, we observe the expected pattern where pLoFs variant groups have the highest proportion associated within each frequency group (Fig. 3B). For high CAF genes and common variants (>1%), this trend is no longer apparent, likely complicated by the presence of artifacts for common pLoF variants (Fig. S19). Second, we compare gene sets by a sampling methodology to match the CAF of gene sets to a background distribution (Fig. 4 and below).

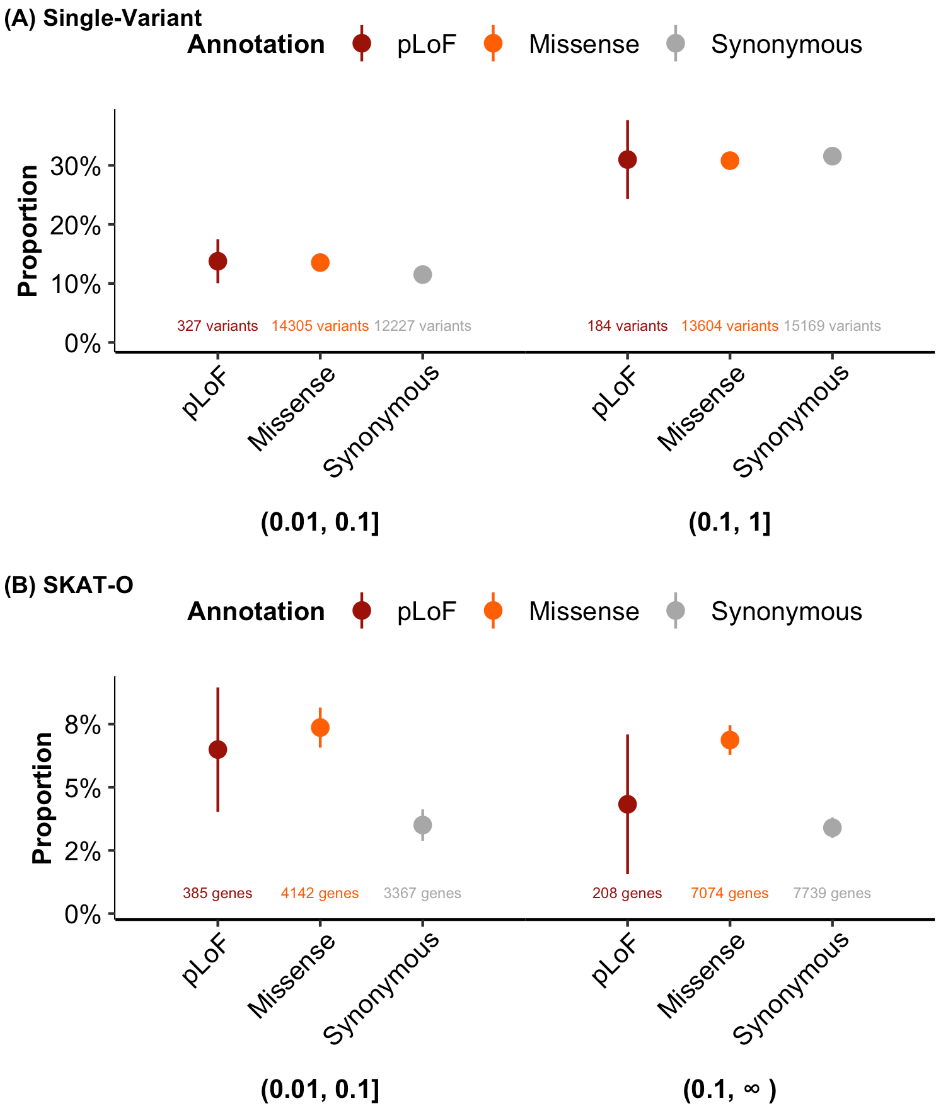


Fig. S19 | The proportion of variants (**A**) and genes (**B**) with at least one phenotype reaching our p-value threshold is shown broken down by allele frequency category (**A**) or cumulative allele frequency category (**B**) and by functional category. For common variants, missense variants show a higher proportion of associations than synonymous variants, but pLoF variants do not show a higher proportion as might be expected, likely due to artifacts at common pLoF variants.

#### Gene set analyses

*Developmental delay genes*

We considered the SKAT-O association results for 470 genes previously implicated in developmental delay (Kaplanis et al., 2020) and compared the number of associations discovered for these genes with the remaining genes in the dataset. To match the background distribution on frequency, we binned genes by their cumulative allele frequency into equal-spaced groups with widths of 0.01, and then matched genes from the remaining set to the distribution of the 470 genes according to their CAF intervals. For each of the three annotation categories (pLoF, missense|LC, and synonymous), we randomly sampled 1,000 matched sets of 470 genes from the remaining set with replacement and computed the mean number of associations and the proportion of genes with at least one association meeting our p-value threshold for each set. By comparing the distribution of the mean and proportion of the 1,000 samples with those of the 470 genes by annotation groups, we found that genes that are implicated in developmental delay are more likely to be associated through a pLoF mechanism with phenotypes in the UK Biobank ($p = 3.6 \times 10^{-4}$, OR = 3.50; Fig. 4).

*Constrained genes*

Similar to the developmental delay genes, we compared 3,917 (1,697 unique genes) constrained gene-annotation pairs from our dataset with the remaining unconstrained genes on their number of associations discovered from SKAT-O results. We obtained LOEUF values for the genes from gnomAD (v2.1.1) and defined constrained genes as those in the highest decile of LOEUF (oe_lof_upper_decile=0). We then matched the unconstrained genes to the constrained genes by CAF intervals with widths of 0.01 and randomly sampled 1,000 unconstrained gene-sets that have sample sizes and CAF distributions comparable to the constrained gene-set for each annotation category with replacement. Finally, we compared the mean number of associations and the proportion of genes with at least one association meeting our p-value threshold of the constrained set with the distribution of the 1,000 unconstrained samples. We found that constrained genes are more likely to be associated with a phenotype in UK Biobank than the unconstrained genes, for pLoF variants ($p = 6.1 \times 10^{-14}$, OR = 4.65; Fig. 4).

*Other gene sets*

We repeat this analysis for a number of gene sets, including autosomal recessive, autosomal dominant, FDA approved drug targets, and GWAS catalog genes, as described in https://github.com/macarthur-lab/gene_lists.

**PolyPhen2 predicted variants**

We compared the proportion of variants with at least one association meeting our p-value threshold across the three PolyPhen2 prediction groups (probably damaging, possibly damaging, and benign; Fig. S20). We binned variants by their minor allele frequency (MAF) into equal-spaced bins with width of 0.01. Using variants from each one of the three groups as reference, we matched the remaining two groups to the reference group by their MAF bins. Relative relationships of the proportion among the three groups are similar when using different reference groups. We then split the variants into allele frequency categories and compared the proportion among different PolyPhen2 prediction groups. We conducted a pairwise proportion test between each pair of groups for each allele frequency interval and observed a significant difference between benign and probably damaging for all three intervals, possibly and probably damaging for allele frequency interval (0.01%, 0.1%] and (1%, 10%], and benign and possibly damaging for allele frequency interval (0.01%, 0.1%] and (0.1%, 1%]. No significant group difference is observed for allele frequencies above 10%.

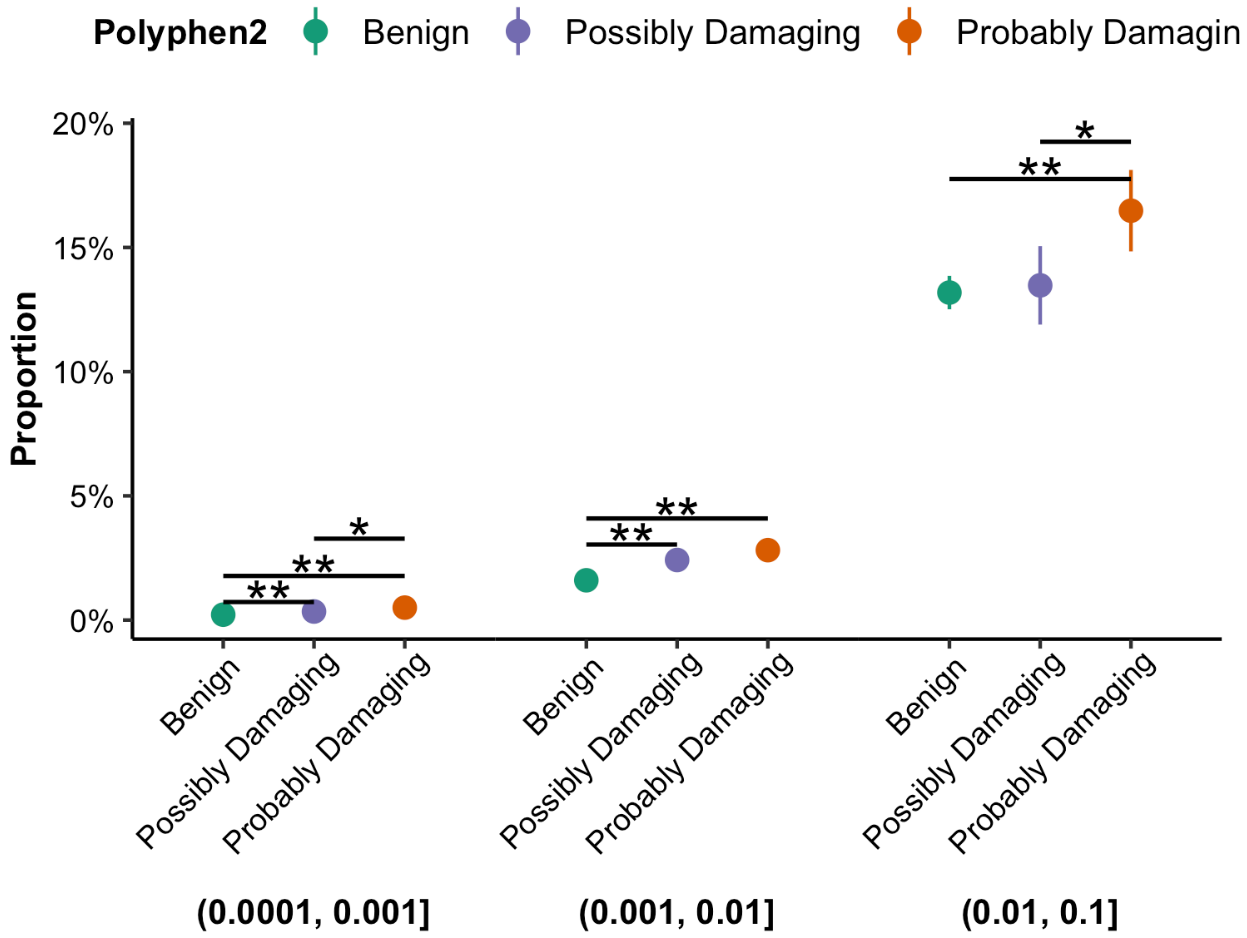


Fig. S20 | The proportion of variants with at least one association is shown broken down by PolyPhen2 annotation group and allele frequency category. * and ** indicate a significant group difference by chi-square test at $p < 0.05$ and $p < 0.001$, respectively. No significant difference is observed for allele frequencies above 10%.

**ClinVar variants**

We obtained pathogenicity of variants from the ClinVar table in gnomAD reference data and then defined pathogenic and likely pathogenic variants as P/LP, benign and likely benign variants as B/LB. We divided the minor allele frequency (MAF) of variants into equal-spaced bins with widths of 0.01 and then matched variants from B/LB and Uncertain significance group to the 1,066 P/LP variants respectively by their MAF bins. For each of the categories, we randomly sampled 1,000 sets matched to the P/LP variant subset with replacement and computed the mean number of associations and the proportion of variants with at least one association meeting our p-value threshold for each set. By comparing the distributions of the mean and proportion of the 1,000 samples with the P/LP group, we found that pathogenic variants are more likely to be associated with a phenotype in the UK Biobank (Fig. 3).

### Data availability and release

Matthew Solomonson, Konrad J. Karczewski, Melissa R. Miller, Ellen Tsai, Nicholas A. Watts, Huy Nguyen, Kevin Nguyen, Cotton Seed, Benjamin M. Neale

**Data availability**

We make the full dataset available in a browser framework (described below), as well as Hail formats, hosted on the Google Cloud Platform. We provide one MatrixTable for each of the single-variant and gene-based tests, as well as Hail Tables with filtering criteria for variants, genes, and phenotypes.

**Code availability**

All code to reproduce the analyses herein is available on github: https://github.com/nealelab/ukb_exomes and https://github.com/nealelab/ukb_common. All quality control and data analysis tasks were performed using Hail (versions between 0.2.49 and 0.2.62) (Hail Team, 2020). Unless otherwise indicated, summarized analyses and plotting were performed in R 4.0.2, using tidyverse.

**Data browser**

Web-based tools like PheWeb have been highly useful in the data processing and dissemination of several recent large-scale biobank genetic studies (Gagliano Taliun et al., 2020; Pruim et al., 2010). PheWeb is well-suited for viewing associations from genotyping data along large genomic regions, where the signal is frequently driven by non-coding regulatory variants rather than variation in protein coding sequences. New web-based tools are needed for visualizing association studies in the context of gene-based analyses. Toward this goal, we previously extended on our gnomAD browser toolkit (Karczewski et al., 2020) to create a suite of portals for displaying gene analysis results from psychiatric exome association studies for schizophrenia (Singh et al., 2020), autism (Satterstrom et al., 2020), bipolar disorder (Palmer et al., 2021), and epilepsy (Epi25 Collaborative, 2019). In this study, we extend our exome browser toolkit to support visualization of biobank-scale PheWAS results. We developed new layouts, navigational mechanisms, plots, and controls that enable users to visualize and compare gene and variant associations across thousands of phenotypes.

*Navigation and workflow*

The browser interface features a novel split-screen design for rapidly inspecting gene-based PheWAS results (Fig. S21). The left hand side displays the global results index, which displays all hits for a given gene, phenotype, or variant (Fig. S21A). The results index displays PheWAS plots or Manhattan plots depending on which navigational button is selected in the top bar (Fig. S21B). The results pane can be condensed, expanded, or hidden entirely by clicking the presets buttons or by dragging the central dotted line left or right. Clicking on one of the arrow buttons in the phenotype table will update the right hand side of the page with a detailed view of the selected gene-phenotype relationship (Fig. S21C). The status bar will update accordingly to reflect the gene, phenotype, variant, or burden dataset that is currently selected (Fig. S21D). Most data are served by a Hail backend, with significant associations cached for speed. Pages load quickly (< 1 second) if the phenotype-gene or phenotype-variant association p-value pair is below the cache threshold ($10^{-4}$ for genes, $10^{-6}$ for variants) or ~4 seconds if above the cache threshold. Partitioning the page in this way allows users to quickly inspect many associations without losing a sense of context, and either half can be easily hidden to create more screen room for information of interest.

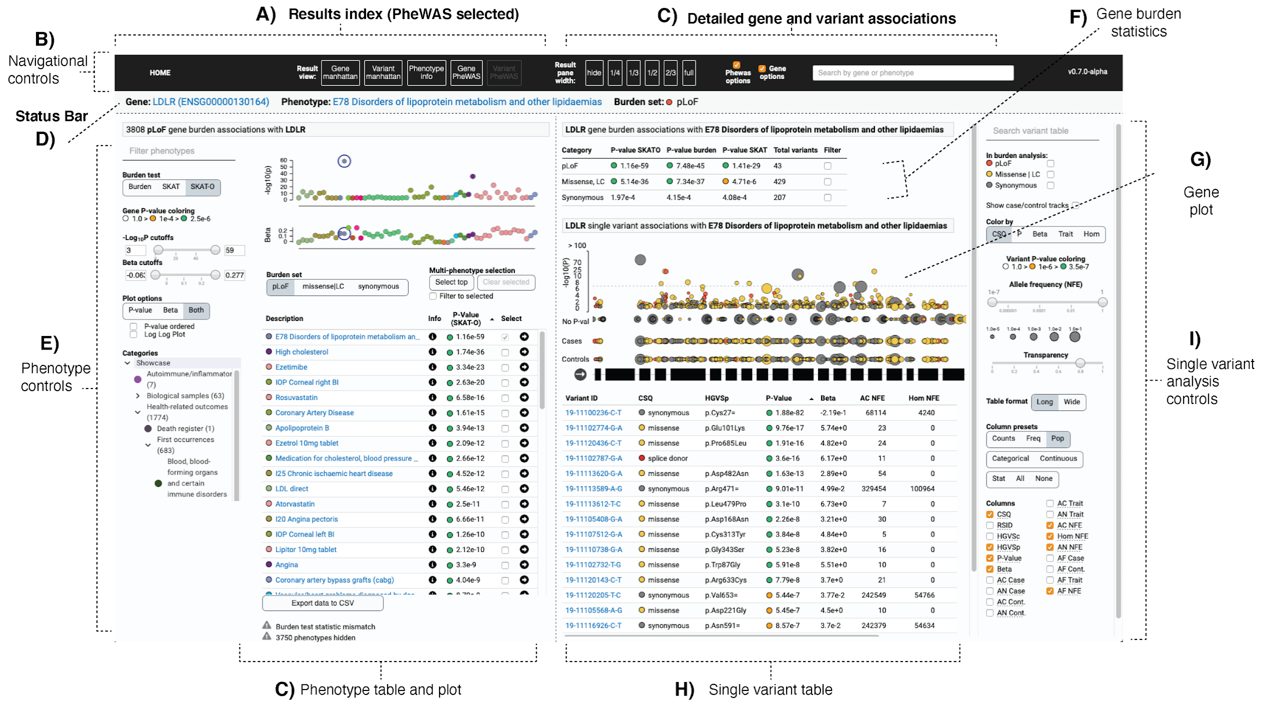


Fig. S21 | Overview of the UKBB exome gene browser interface. The left hand side of the page provides access to all associations with a given gene, variant, or phenotype. The right hand side is for exploring detailed gene test associations (burden, SKAT-O, SKAT) across annotation groups (pLoF, missense and low confidence pLoF, synonymous) in addition to single variants that were included in the burden tests.

*Exploring associations by gene*

When the “gene PheWAS” results pane is active, the results index displays all phenotypes associated with a particular gene in PheWAS plot and tabular formats (Fig. S21C). The phenotype control panel (Fig. S21E) enables users to specify which of the three burden tests (Burden, SKAT, SKAT-O) or mutational class (pLoF, missense, synonymous) test statistics to display. Phenotypes can be filtered by keywords such as phenotype description or trait type (continuous, categorical, or ICD10). The results can also be filtered by p-value or beta using minimum and/or maximum thresholds. Note that for genes, the beta statistic is always derived from the burden test (SKAT and SKAT-O do not produce beta statistics). The PheWAS plot is colored and grouped by UK biobank showcase category; the category control section can be used to traverse the showcase tree and filter the phenotype list to those belonging to specific categories. The PheWAS plot can be configured to show p-values on either log or double log scales. Users can expand the plot to focus on p-values only, betas-only, or view p-values and betas simultaneously.

The gene burden statistics table summarizes burden results across all mutational classes and tests (Fig. S21F). The gene plot displays single variants mapped to genomic coordinates along the gene exons. Variant $-\log_{10}p$ values are shown on the Y axis (Fig. S21G). The plot transitions from a single log scale to a double log scale ⅔ along the plot height to prevent variants with extremely low P-values from dominating the plot, allowing users to focus on novel rare variant associations near the significance threshold. Variants are depicted as circles, with the circle radii log-scaled by allele frequency in the non-Finnish European population. By default, variants are colored by their most severe VEP consequence across transcripts. If the selected phenotype is categorical, two additional case/control variant tracks display variant positions with radii log-scaled to allele frequencies in cases and controls, respectively. If the selected phenotype is continuous, variant radii will be log-scaled by allele frequency among individuals measured for the trait.

The single variant analysis control panel is used to configure data displayed related to single variants (Fig. S21I). Checkboxes enable users to filter variants to those included in the gene burden analysis. Variants are filterable by identifier or annotation using the search box. Users can focus on particular parts of the allele frequency spectrum by dragging the allele frequency filter slider. Detailed summary statistics for all exome variants are available in a table below the plot (Fig. S21H). Users can specify which columns to display using the column selection checkboxes, or they can choose one of the column group presets. Each preset will select a particular set of columns that make sense to compare side-by-side (e.g., allele counts, frequencies, population counts, and columns best suited for categorical or continuous trait types).

*Exploring associations by phenotype*

When the “gene Manhattan” results pane is active, the results index displays all gene associations with a selected phenotype. The results are displayed in Manhattan plot, QQ plot, and tabular format (Fig. S22). The three burden test types are displayed as columns, and the burden set (pLoF, missense|LC, or synonymous) can be selected with the “Burden set” segmented control. Clicking on a gene name will navigate to the gene PheWAS view, and clicking on the “details” arrow will update the right hand side without leaving the gene Manhattan view. When the “variant Manhattan" results pane is active, the left hand side results index takes a similar format as the gene Manhattan but displays single variant association p-values instead of gene test statistics. Single variant results can be filtered by consequence category (pLoF, missense, synonymous, and other). Clicking a variant ID will navigate to that variant’s PheWAS view, and clicking the “details” arrow will keep the single variant Manhattan view active.

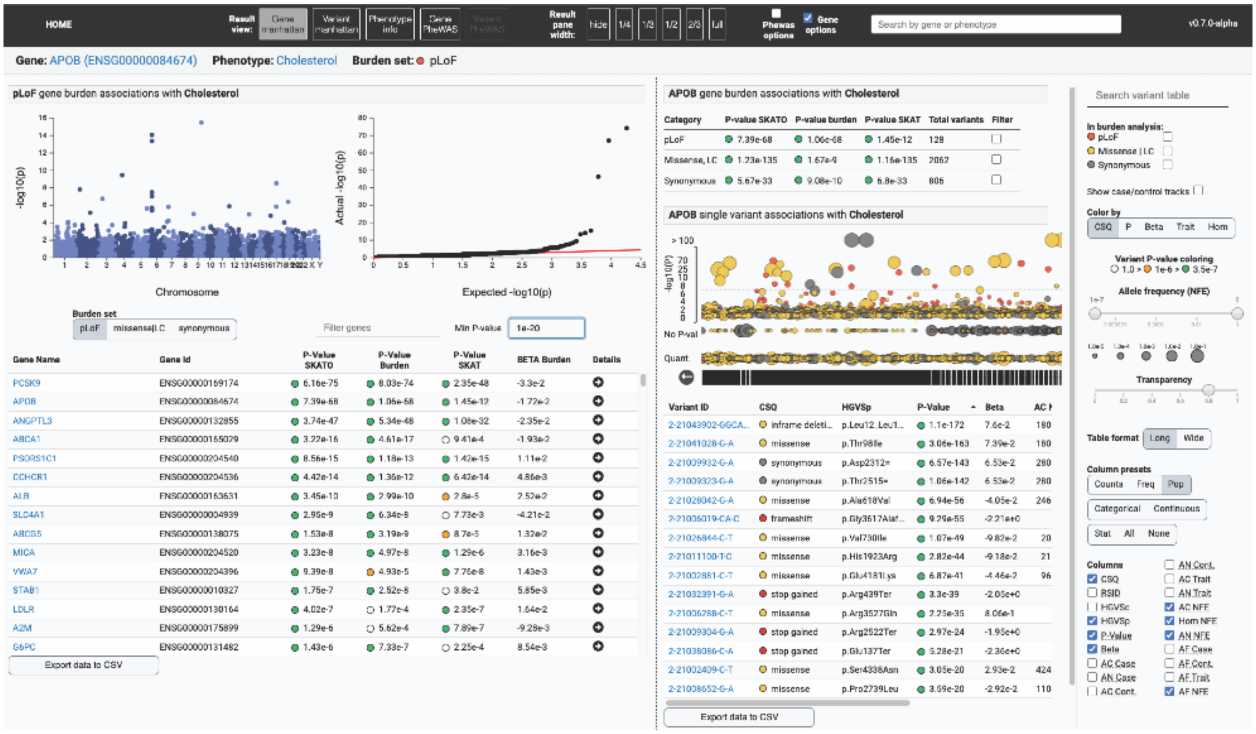


Fig. S22 | Results by phenotype. For a given phenotype, gene or variant association results are displayed in Manhattan plot formats in addition to an exportable table. Detailed gene results can be quickly previewed using the arrow button located in each row of the table.

*Comparing single variant associations across phenotypes*

For a more comprehensive view of all variant-level associations for a gene in a single view, we developed functionality for exploring many phenotypes simultaneously on the gene page (Fig. S23). This feature aims to help users gain insight into pleiotropic patterns of variation across all high-scoring traits for a gene. Each row in the PheWAS table has a checkbox that, when checked, will overlay the phenotype in the gene plot (Fig. S23A). The “select top” button will load all phenotypes below the $10^{-4}$ p-value threshold; the “clear selected” button will unselect all phenotypes and return to the single phenotype view (Fig. S23B). When selected, phenotypes are assigned randomly generated colors to make them easier to distinguish in the plot and table (Fig. S23C). Many tens or hundreds of phenotypes can be loaded simultaneously; however, an automatic p-value threshold will be applied when there are too many variants to display and the user will be warned in the single variant control panel.

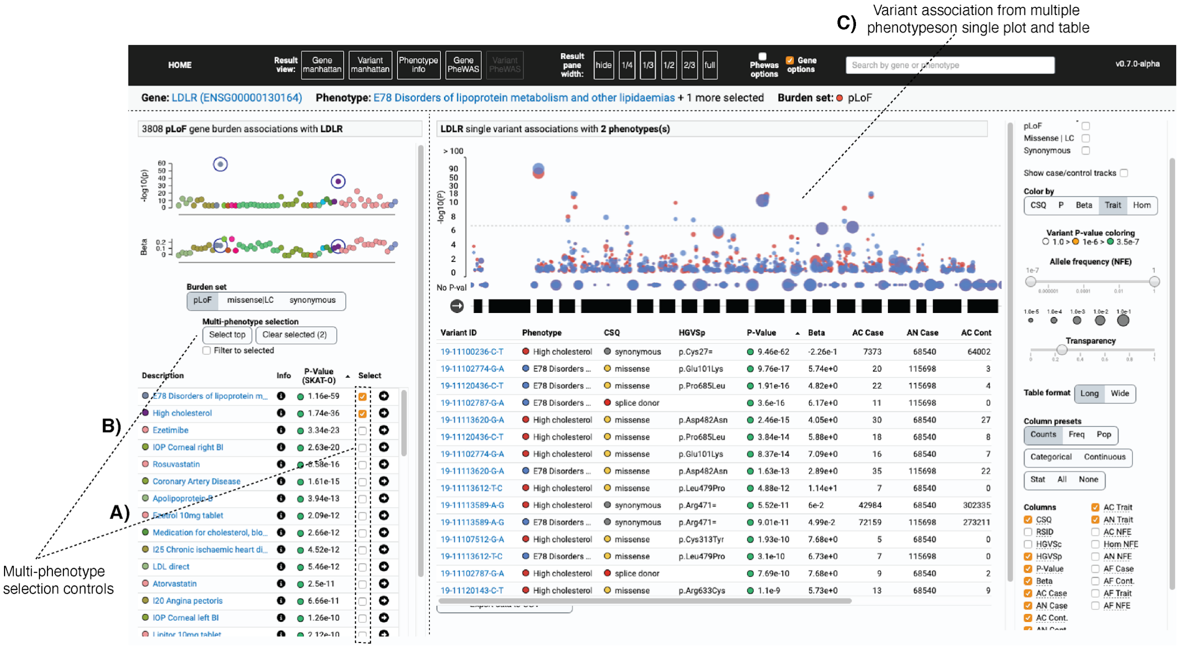


Fig. S23 | Multi-phenotype plotting. Many phenotypes can be selected simultaneously to be overlaid for comparison of single variant analysis associations.

By default, the variant table is configured to the “long” table format; when multiple phenotypes are selected, each variant-phenotype association will appear as a row in the variant table such that the variant table now contains duplicate entries for each variant. To make rows unique and to see comparison of association statistics across traits in a single row, the table can be set to “wide” format (Fig. S24A). The phenotype pivots to the columns, creating a sort of genotype-phenotype matrix (Fig. S24B). The column selection controls will affect both the long and wide table formats. When examining many phenotypes at once, users can click the “filter to selected” button on the phenotype section to simplify the PheWAS plot by only showing the selected phenotypes; this effectively serves as a legend for coloring-by-trait functionality (Fig. S24C).

Hover interactions are especially useful when comparing multiple phenotypes; hovering over variants or phenotypes with the mouse will emphasize the relevant variants and bring them to the foreground (Fig. S24D). The transparency slider sets the opacity level for non-hovered variants, helping the user tune the multi-phenotype plot such that hovered selections can stand out better (Fig. S24E).

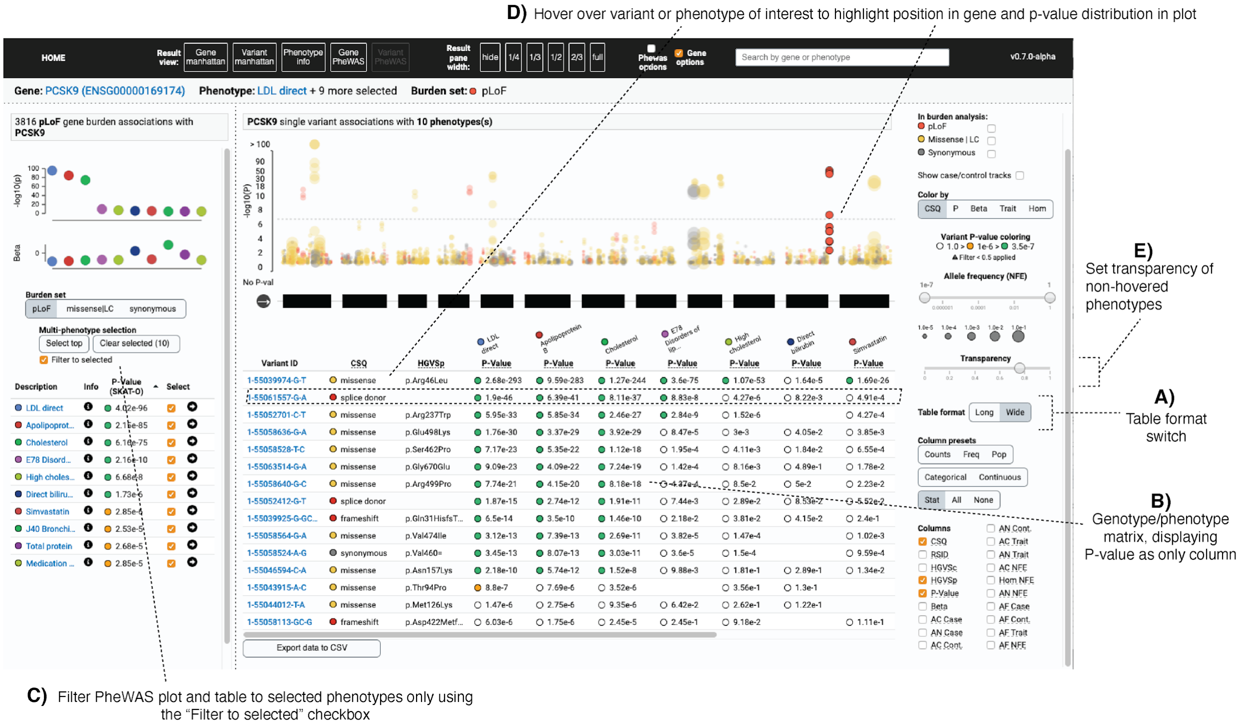


Fig. S24 | Using hover interactions with the multi-phenotype pivot table. Here 10 LDLR associations are compared simultaneously and one splice donor of interest is hovered in the variant table to highlight the plot.

For categorical traits in particular, it is useful to get a visual sense of how case/control variant positions and allele frequencies differ across the gene. The "show case/control tracks" checkbox will fold out case/control tracks for all traits currently selected (Fig. S25A). Continuous traits will be displayed in a single track and the allele frequency for individuals measured for the trait will be displayed. By viewing the case/control counts in the study (Fig. S25B), alongside the case/control allele counts for variants in the variant table (Fig. S25C) and the plots (Fig. S25D) users can very quickly compare burden results across phenotypes, genes, and individual variants to get a sense of which specific variants may be driving gene burden signals. When the per-phenotype tracks are expended, it can be useful to use the "Color by" switch to look for trends and variants across genomic coordinate, trait, consequences, association p-value, effect size, and zygosity (Fig. S26).

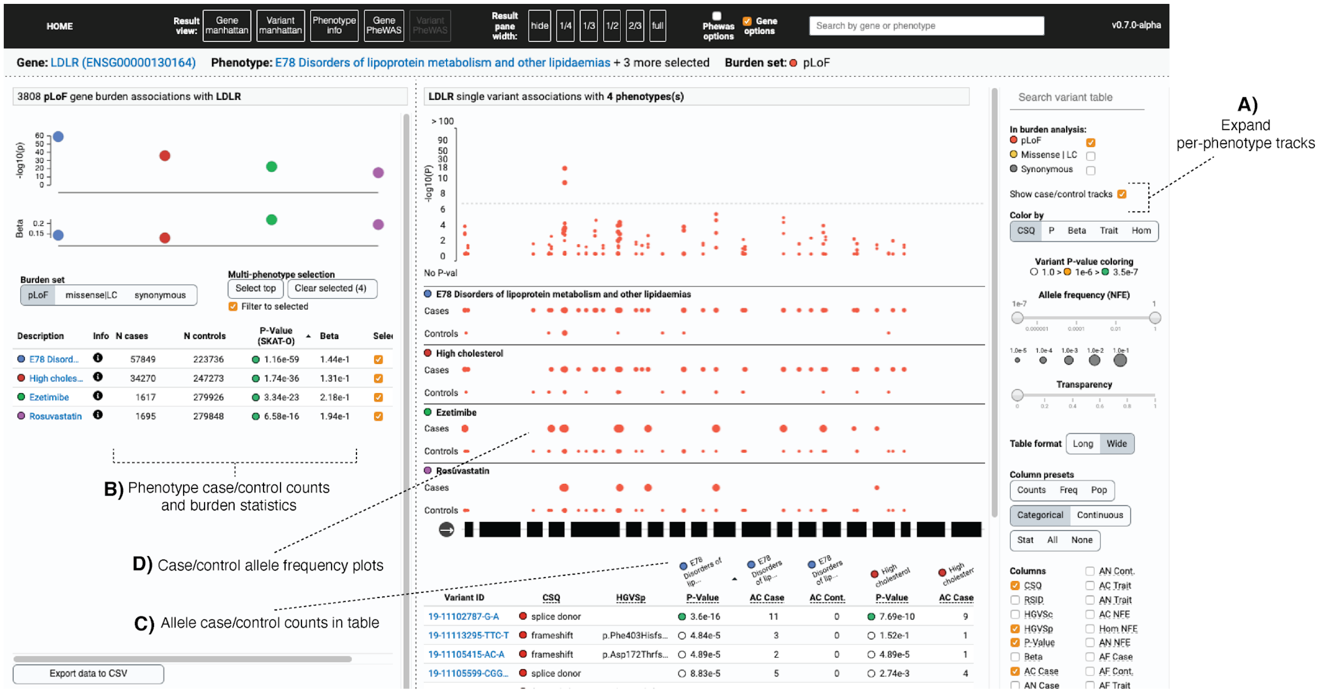


Fig. S25 | Viewing case-control counts and allele frequencies for pLoF variants across traits in a gene.

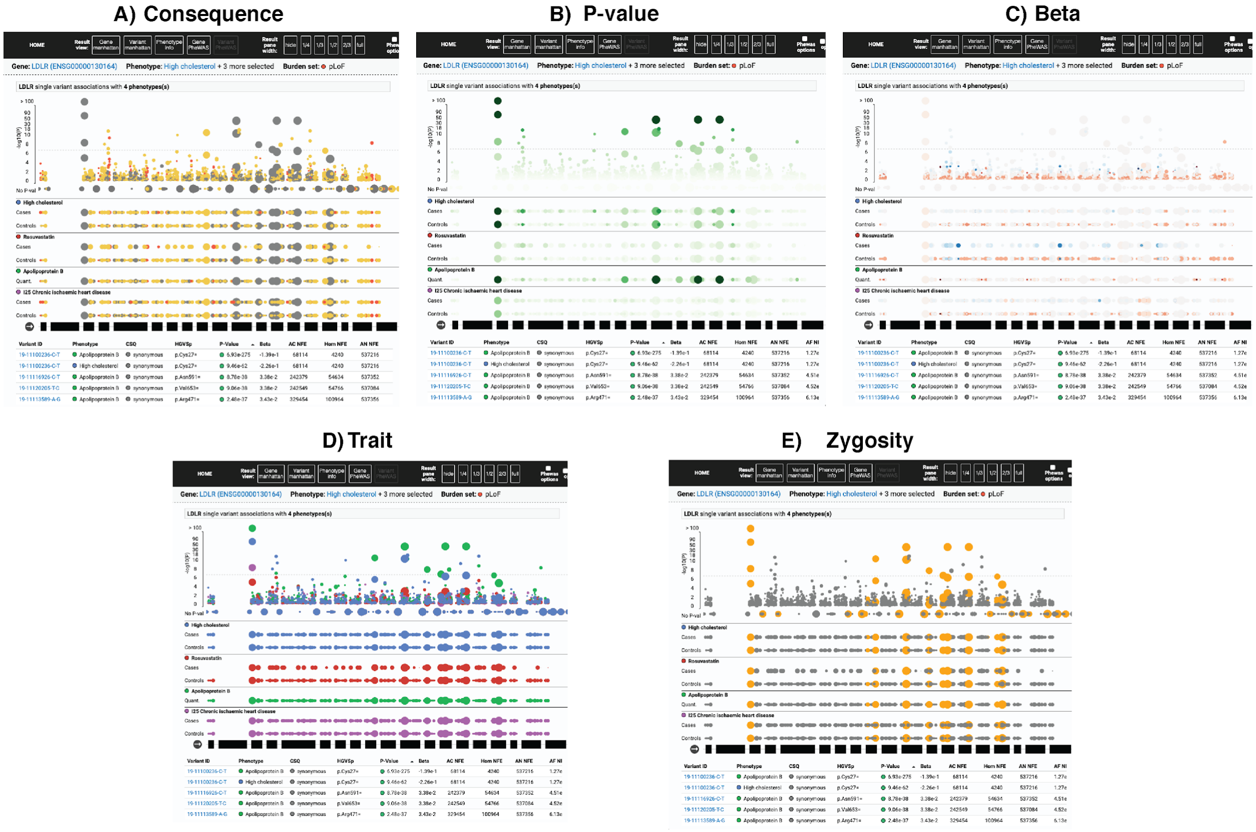


Fig. S26 | Color variants by attribute to uncover patterns in A) consequence, B) p-value, C) beta, D) trait, or E) zygosity.

*Single variant results*

When a variant is clicked on the single variant Manhattan plot or on the gene page, the page will focus on the selected variant, and a PheWAS displays all associations with that variant (Fig. S27). Similar to the gene PheWAS page, multiple phenotypes can be selected and loaded at once. In the variant view, the table rows show statistics for the selected variant, and the columns show values across selected phenotypes. The variant position is displayed along the genomic coordinate. Clicking the “unselect” button will return to the gene page. In this way, users can easily flip back and forth between single variants and the gene context.

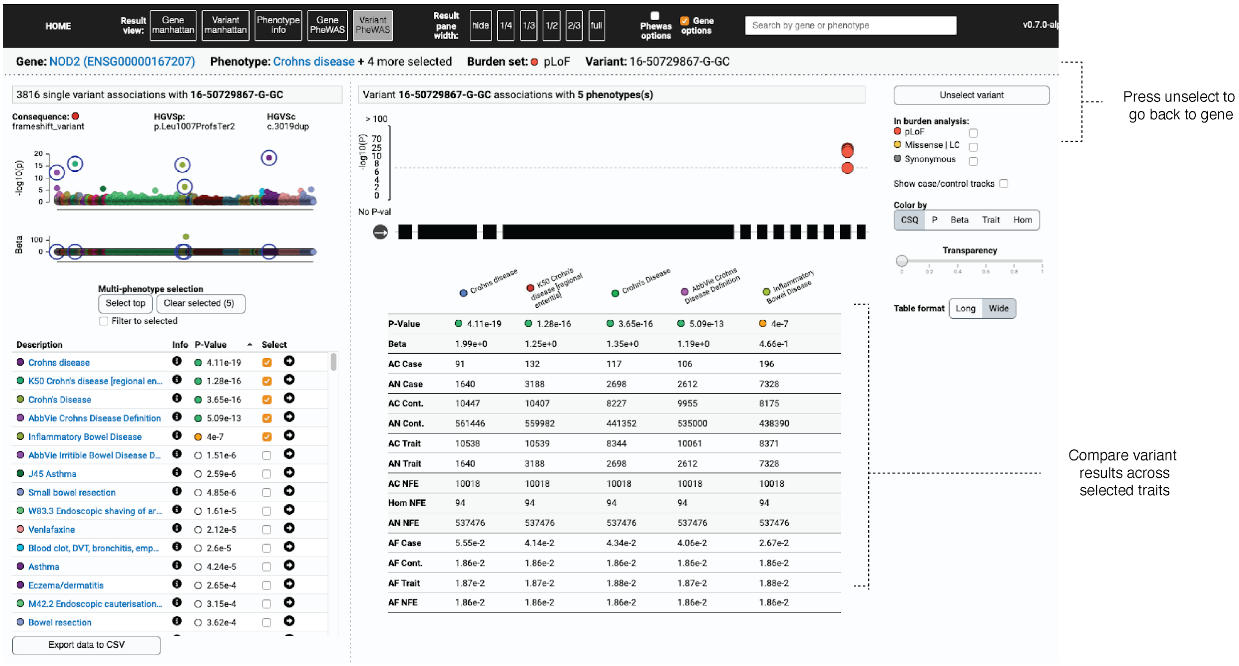


Fig. S27 **|** Single variant page.
